# Multi-country and intersectoral assessment of cluster congruence between different bioinformatics pipelines for genomics surveillance of foodborne bacterial pathogens

**DOI:** 10.1101/2024.07.24.24310933

**Authors:** Verónica Mixão, Miguel Pinto, Holger Brendebach, Daniel Sobral, João Dourado Santos, Nicolas Radomski, Anne Sophie Majgaard Uldall, Arkadiusz Bomba, Michael Pietsch, Andrea Bucciacchio, Andrea de Ruvo, Pierluigi Castelli, Ewelina Iwan, Sandra Simon, Claudia E. Coipan, Jörg Linde, Liljana Petrovska, Rolf Sommer Kaas, Katrine Grimstrup Joensen, Sofie Holtsmark Nielsen, Kristoffer Kiil, Karin Lagesen, Adriano Di Pasquale, João Paulo Gomes, Carlus Deneke, Simon H. Tausch, Vítor Borges

## Abstract

Food and waterborne disease (FWD) surveillance requires Whole-Genome Sequencing (WGS)-based systems following a One Health approach. However, different laboratories employ different WGS pipelines in their routine surveillance activities, casting doubt on the comparability of their results and hindering optimal communication at intersectoral and international levels. Through a collaborative effort involving eleven European institutes across seven countries and spanning the food, animal and human health sectors, we aimed to assess the inter-laboratory comparability of WGS clustering results for four important foodborne pathogens: *Listeria monocytogenes*, *Salmonella enterica*, *Escherichia coli* and *Campylobacter jejuni*. Each participating institute (n=9) applied its surveillance pipeline over the same WGS datasets (>2000 isolates per species), and, for each pipeline, genetic clusters were identified at each possible allele/SNP distance threshold. Inter-pipeline clustering congruence was assessed by calculating a “Congruence Score” (relying on Adjusted Wallace and Adjusted Rand coefficients) across all resolution levels, followed by an in-depth comparative analysis of cluster composition at outbreak level. An additional cluster congruence assessment was performed between WGS and traditional typing, which, depending on the species, included Sequence Type (ST), Clonal Complex (CC) and/or serotype. Our results revealed a general high concordance between allele-based pipelines at all resolution levels for all species, except for *C. jejuni*, where the different resolution power of available allele-based schemas led to marked discrepancies. Still, this study identified non-negligible differences in allele-based pipeline performance for outbreak cluster detection, suggesting that a threshold flexibilization is important for the detection of similar outbreak signals by different laboratories. These results, together with the observation that different STs, CCs and serotypes exhibit remarkably different genetic diversity, should inform future threshold selections for outbreak case definitions. In conclusion, this study provides valuable insights into the comparability of pipelines commonly used for routine genomics surveillance, and reinforces the need, while demonstrating the feasibility, of conducting continuous and comprehensive WGS pipeline comparability assessments. Ultimately, it opens good perspectives for a smoother international and intersectoral cooperation and communication towards a sustainable and efficient One Health FWD surveillance.

## Introduction

Food and waterborne diseases (FWDs) affect 600 million people every year worldwide and represent an important burden for human and animal health ^1^. Therefore, FWDs prevention and control is of high importance, requiring adequate surveillance systems able to track the circulation of pathogens, detect and investigate potential outbreaks, and monitor their clinical and epidemiological relevant features ^2,3^. Such systems must recognize the interconnection between people, animals, plants, and their shared environment, following a One Health approach ^2,4,5^.

International agencies, such as the World Health Organization (WHO), the World Organization of Animal Health (WOHA), the European Center for Disease Prevention and Control (ECDC) and the European Food Safety Authority (EFSA), strongly promote the integration of Whole-Genome Sequencing (WGS) data as an essential component of surveillance systems ^6–15^. Therefore, a significant effort is in place to develop and refine surveillance-oriented bioinformatics solutions for the assessment of samples’ genomic relatedness and outbreak cluster identification. These include automated command-line pipelines, such as SnapperDB^16^, chewieSnake ^17^ or WGSBAC ^18,19^, user-friendly platforms, like INNUENDO ^20^, BigsDB/PubMLST ^21^, IRIDA ^22^, PathoGenWatch ^23^, COHESIVE ^24,25^ or Enterobase ^26^, and also commercial software, as Bionumerics or Ridom SeqSphere+ ^8,27^. Although these solutions cover the basic steps of a WGS data analysis pipeline (and some species-specific requirements), each of them has its own specificities and particularities ^8^.

From a technical perspective, genomic pipelines can be roughly divided into allele- and SNP-based pipelines, where genomic relatedness is either assessed based on the alleles present at a given set of loci (schema), or on the comparison of their single nucleotide polymorphisms (SNPs). Allele-based (also known as gene-by-gene) pipelines may rely on a core-genome Multilocus Sequence Type (cgMLST) approach, in which the schema corresponds to a pre-defined group of loci expected to be present in most of the species’ isolates, or, alternatively, in a whole-genome Multilocus Sequence Type (wgMLST) approach, in which the schema includes both the core and accessory loci, possibly providing a higher resolution power ^27,28^. On the other hand, SNP-based pipelines commonly rely on the alignment of sequencing reads to a closely related reference genome, but *k-*mer- and assembly-based alignments are also available as alternatives to detect SNPs ^27,29^. In the specific case of foodborne bacterial pathogens, although SNP-based pipelines can be used as a first-line approach to detect potential outbreak-related clusters, they are more often used for fine-tuned analyses with a small set of samples at high-resolution levels in order to confirm their genomic proximity as inferred by an cg/wgMLST approach ^16,27–30^. Indeed, nowadays, allele-based pipelines are promoted internationally as the first-line approach for monitoring pathogen populations and detection of clusters of samples related to potential outbreaks ^6–8,27,31^, being the most commonly used at European level, as demonstrated by recent ECDC “External Quality Assessments” (EQAs) ^32–34^.

European laboratories have been following independent paths towards the implementation of FWD genomic surveillance frameworks, often being at different stages of this technological transition and ending up with different bioinformatic solutions for outbreak cluster detection ^2,32–35^. This heterogeneity raises concerns regarding inter-laboratorial communication of surveillance results, with special relevance during multi-country outbreak investigations where WGS-based criteria for case definition is often similar regardless of the pipeline ^36–39^. Although previous studies have found some comparability in the clustering results at outbreak level obtained by different pipelines ^40–42^, and public health authorities routinely launch international EQAs ^43,44^, there is still a need for large-scale studies providing in-depth knowledge about the congruence of the routinely applied WGS strategies. Such studies would be aligned with international agencies guidelines, which warn of the need to ensure the harmonization and comparability of outputs resulting from different methods ^5,13,14^.

In the frame of the BeONE project of the “One Health European Joint Programme” (OHEJP), eleven Institutes across Europe, spanning different sectors, cooperated to advance in the field of FWD surveillance ^45^. This project aimed to contribute to the capacity of European laboratories to routinely integrate genomic and epidemiological data, and facilitate data sharing and comparability among EU countries, international organizations, and/or other stakeholders involved in FWD prevention and control. In the present study, we (the BeONE consortium) assessed the congruence and comparability of the clustering results obtained through the various WGS bioinformatics pipelines used by the consortium partners for genomics surveillance of four important foodborne bacterial pathogens (*Listeria monocytogenes, Salmonella enterica, Escherichia coli* and *Campylobacter jejuni*). This is a crucial step to promote efficient communication at international and intersectoral levels towards the establishment of a fully integrative One Health genomic surveillance framework, according to the best practices and recommendations ^5,13,14,46^.

## Results

### 1. Study design and strategy for congruence analysis

Multiple European laboratories, from different countries and sectors, employed, whenever possible, their different bioinformatics pipelines for genomic surveillance of foodborne bacterial pathogens (Figure 1) on four WGS datasets of *L. monocytogenes* (n = 3300 isolates ^47,48^), *S. enterica* (n = 2974 isolates ^49,50^), *E. coli* (n = 2307 isolates ^51,52^), and *C. jejuni* (n = 3686 isolates ^53,54^) ([Additional file](https://zenodo.org/records/12805751) 1, details in the Methods section). After Quality Control (QC), allele-based pipelines ran with the whole datasets, while SNP-based independently ran with sub-datasets of the top represented Sequence Types (STs) or serotypes of each species (Table 1 and [Additional file](https://zenodo.org/records/12805751) 1) to better mimic their common application in the discrimination of closely related strains (e.g. same ST or outbreak-related strains). This collaborative effort covered a broad variety of “pipelines” (hereinafter used as a proxy for the combination of software pipeline and schema/reference), including the most commonly used cg/wgMLST schemas, allele/SNP-callers and clustering methods (Table 1).

**Figure 1.**
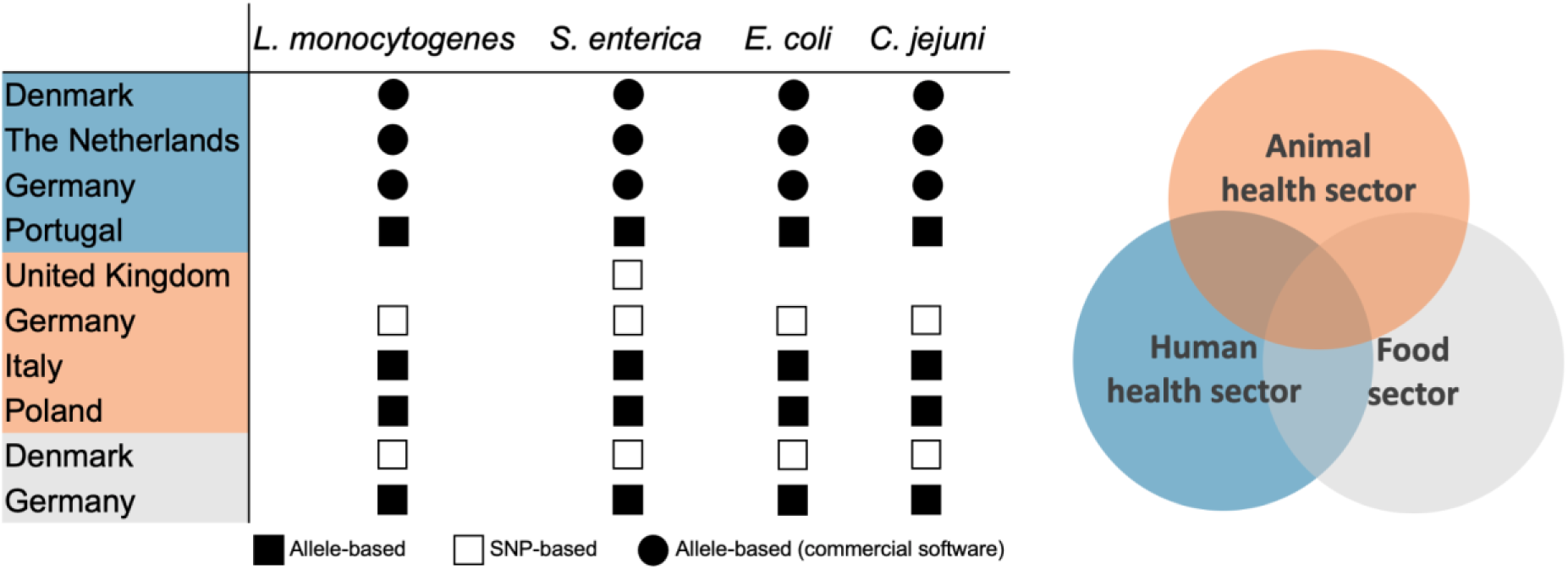
Summary of the different countries and sectors involved in the pipeline cluster congruence assessment, with indication of the diversity of pipelines used for FWD surveillance per country, sector and species.

**Table 1.**
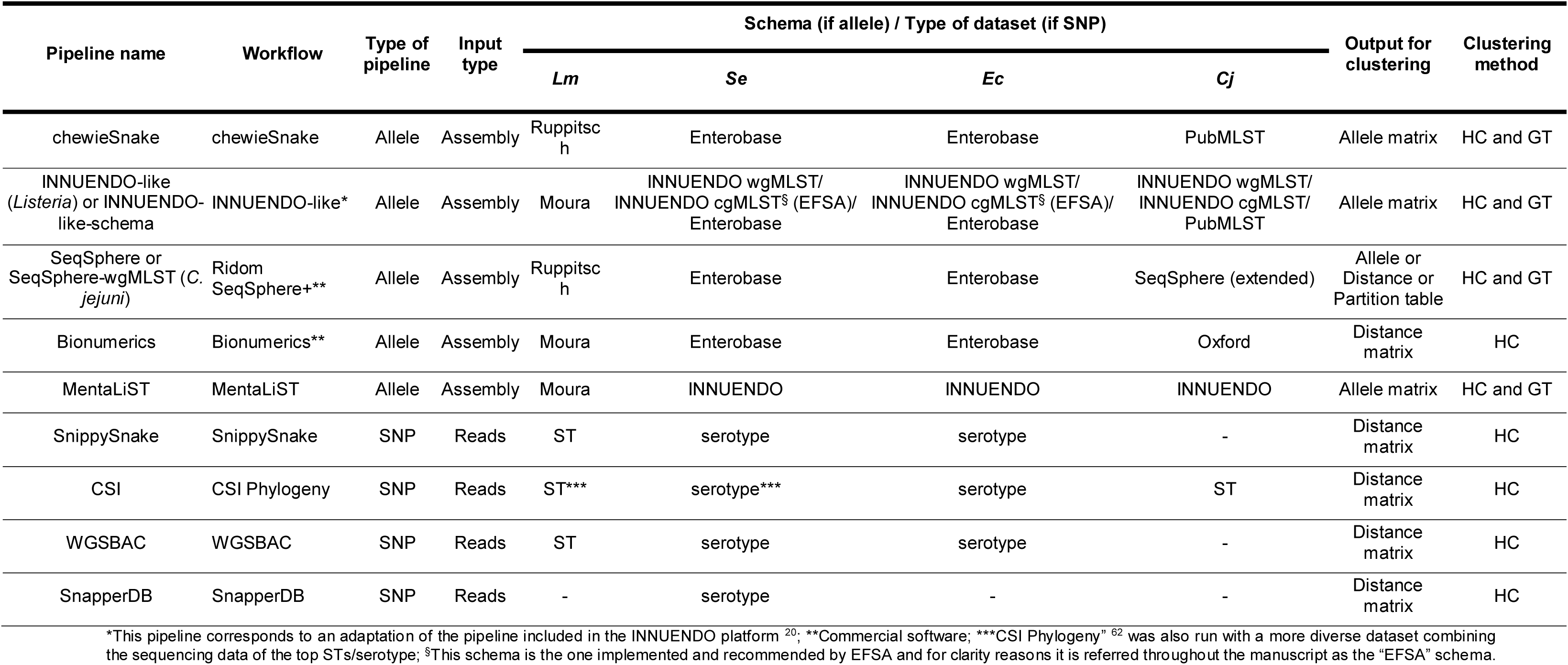
Pipelines used for the cluster congruence analysis with indication of the input type, the schema or reference genome used for each species, the output used for clustering and the clustering method(s) applied. HC – Hierarchical clustering; GT – GrapeTree.

| Pipeline name | Workflow | Type of pipeline | Input type | Schema (if allele) / Type of dataset (if SNP) |  |  |  | Output for clustering | Clustering method |
| --- | --- | --- | --- | --- | --- | --- | --- | --- | --- |
|  |  |  |  | *Lm* | *Se* | *Ec* | *Cj* |  |  |
| chewieSnake | chewieSnake | Allele | Assembly | Ruppitsch | Enterobase | Enterobase | PubMLST | Allele matrix | HC and GT |
| INNUENDO-like (*Listeria*) or INNUENDO-like-schema | INNUENDO-like* | Allele | Assembly | Moura | INNUENDO wgMLST/<br>INNUENDO cgMLST<sup>§</sup> (EFSA)/<br>Enterobase | INNUENDO wgMLST/<br>INNUENDO cgMLST<sup>§</sup> (EFSA)/<br>Enterobase | INNUENDO wgMLST/<br>INNUENDO cgMLST/<br>PubMLST | Allele matrix | HC and GT |
| SeqSphere or SeqSphere-wgMLST (*C. jejuni*) | Ridom SeqSphere+** | Allele | Assembly | Ruppitsch | Enterobase | Enterobase | SeqSphere (extended) | Allele or Distance or Partition table | HC and GT |
| Bionumerics | Bionumerics** | Allele | Assembly | Moura | Enterobase | Enterobase | Oxford | Distance matrix | HC |
| MentaLiST | MentaLiST | Allele | Assembly | Moura | INNUENDO | INNUENDO | INNUENDO | Allele matrix | HC and GT |
| SnippySnake | SnippySnake | SNP | Reads | ST | serotype | serotype | - | Distance matrix | HC |
| CSI | CSI Phylogeny | SNP | Reads | ST*** | serotype*** | serotype | ST | Distance matrix | HC |
| WGSBAC | WGSBAC | SNP | Reads | ST | serotype | serotype | - | Distance matrix | HC |
| SnapperDB | SnapperDB | SNP | Reads | - | serotype | - | - | Distance matrix | HC |

\*This pipeline corresponds to an adaptation of the pipeline included in the INNUENDO platform <sup>20</sup>; \*\*Commercial software; \*\*\*CSI Phylogeny” <sup>62</sup> was also run with a more diverse dataset combining the sequencing data of the top STs/serotype; <sup>§</sup>This schema is the one implemented and recommended by EFSA and for clarity reasons it is referred throughout the manuscript as the “EFSA” schema.

In order to assess pipeline clustering congruence and comparability, it was essential to obtain clustering information at all possible distance thresholds for each pipeline (both allele- and SNP-based pipelines). Given the heterogeneity of “end-point” pipeline outputs, this harmonization was achieved by ReporTree ^55^, which also allowed the application of the clustering methods used by the original laboratory, either single-linkage hierarchical clustering (HC) or Minimum-Spanning Tree (MST) generation through MSTreeV2 model of GrapeTree (GT) ^56^ (Table 1, details in the Methods section). In addition, whenever an allele matrix was provided, clustering was performed with both algorithms, reinforcing the power and magnitude of the comparison (Table 1). We took advantage of this vast amount of data to conduct four relevant complementary analyses:

#### i. Evaluation of allele-based clustering and comparison of stability regions

This preliminary evaluation targeted the pipelines running the whole dataset (i.e. allele-based pipelines) and aimed at providing a global perspective on the resolution power of each pipeline and identifying ranges of allelic distance (AD) thresholds associated with cluster stability, i.e., subsequent thresholds in which clustering remains similar (potentially relevant for nomenclature design). For this, we assessed the number and composition of clusters obtained at progressively increasing distance thresholds and determined the neighborhood Adjusted Wallace coefficient (nAWC), which varies from 0 (no congruence) to 1 (absolute congruence), at consecutive thresholds (“n + 1” → “n”), based on a previously described approach ^57–59^.

#### ii. Evaluation of allele-based clustering congruence with traditional typing data

This assessment targeted the pipelines running the whole dataset (i.e. allele-based pipelines) and aimed at identifying the threshold levels (and stability regions) with the highest congruence with traditional typing (ST, Clonal Complex [CC] or serotype) and, when possible, with WGS-derived pathogen main lineages. To this end, for each comparison, we calculated the AWC between each threshold of the allele-based typing and these classifications, as a measure of the probability that two samples that cluster together using one method (at a given threshold level) also belong to the same lineage, ST, CC or serotype 58. This was conducted between all possible threshold levels in both directions (method A → method B and method B → method A). In addition, for each comparison, we also calculated the Adjusted Rand (AR) coefficient as a measure of the overall agreement between the typing methods ^58^. The three values calculated for each comparison were then combined into a “Congruence Score” (CS) (CS = AWC_A → B_ + AWC_B → A_ + AR), which varies from 0 (no congruence) to 3 (absolute congruence). By providing a more intuitive interpretation of WGS-based typing outputs, this evaluation is expected to show how the cg/wgMLST clustering relates with historical typing data, thus facilitating the adoption of WGS-based surveillance by laboratories starting this technological transition.

#### iii. Evaluation of cluster congruence between different pipelines at all threshold levels

This evaluation was conducted for the whole dataset (involving the pairwise comparison of all allele-based pipelines) and for the sub-datasets of the top represented STs or serotypes of each species (involving the pairwise comparison of both allele- and SNP-based pipelines). We aimed to determine: i) the threshold levels with the highest concordance between the different pipelines (i.e., threshold levels/regions with similar clustering); ii) whether the same clustering results are obtained at similar threshold levels for both pipelines; and, iii) the existence of regions of stability common to all pipelines. For each pairwise pipeline comparison, we assessed the above-mentioned coefficients and CS through the comparison of all thresholds from the lowest to the highest resolution, and identified the thresholds with highest concordance between two pipelines (“corresponding points”). These comparisons provide key information to facilitate international data sharing and cooperation, and to smooth future workflow upgrades (either to accommodate technological advances or upcoming international recommendations).

#### iv. Evaluation of pipeline performance in identifying potential outbreak-related clusters

This analysis aimed at assessing congruence and threshold variability to detect the same potential outbreak-related clusters. For this, we identified the genetic clusters at potential outbreak-level using each allele-based pipeline and, subsequently, assessed the thresholds at which the same cluster composition was (if it was) found by the other ones. By targeting the main focus of routine surveillance (detection and survey of clusters of potential public health interest), this output and associated tools are expected to facilitate and increase confidence in the direct communication between laboratories during a real-life outbreak scenario.

### 2. Listeria monocytogenes

*Listeria monocytogenes* dataset (Figure 2A) was analyzed with five allele-based and three SNP-based pipelines (Table 1 and Tables S1.1 and S1.2 in Additional file 2).

**Figure 2.**
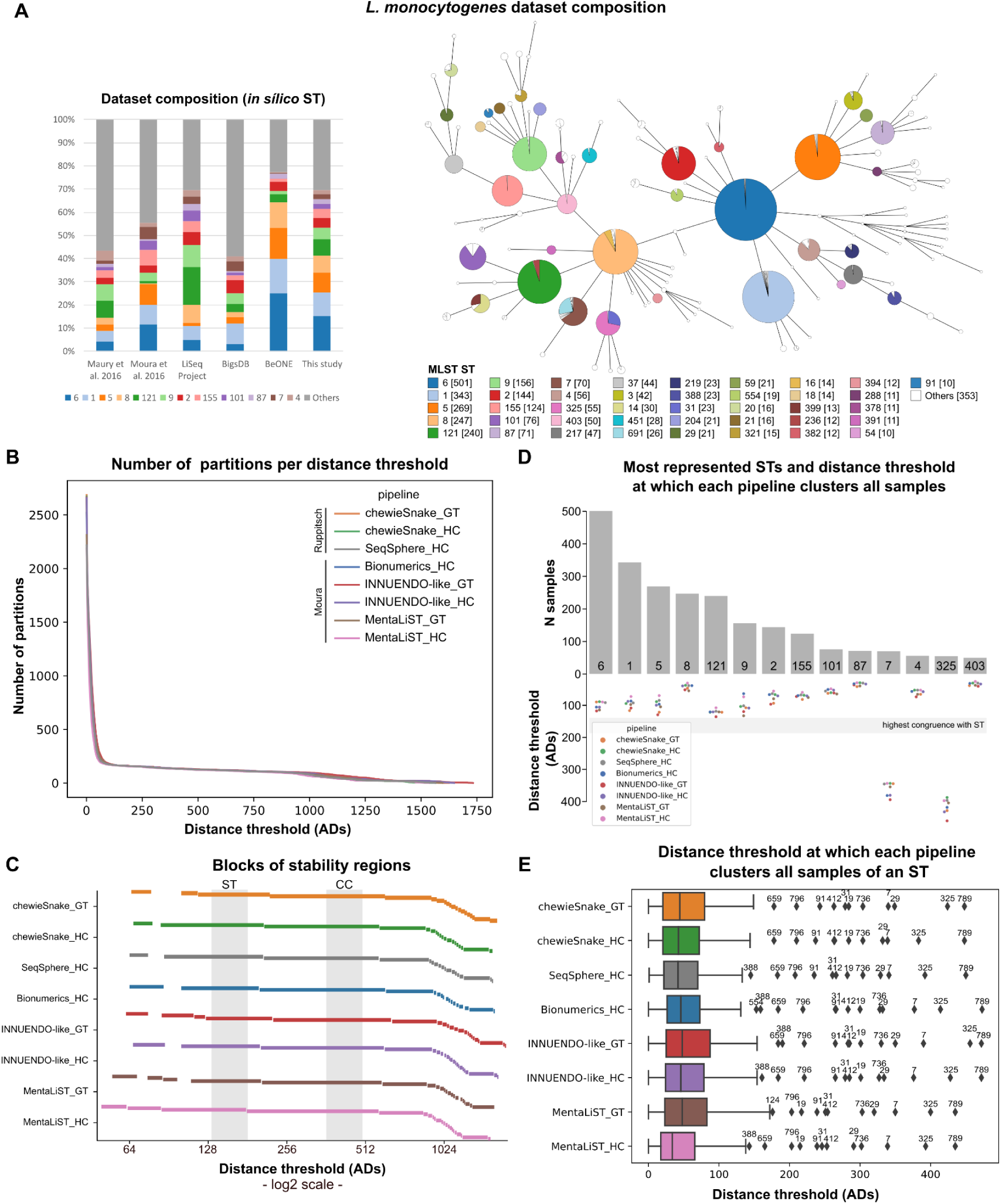
Assessment of allele-based clustering at all possible threshold levels for *L. monocytogenes* and congruence with traditional MLST. **A)** Composition of the *L. monocytogenes* dataset used in this study in terms of ST in comparison with datasets of previous studies (Maury et al. 2016 ^127^ and Moura et al. 2016 ^60^), the LiSeq project ^128^ and the BIGSdb database as of November 2021 ^21^. A GrapeTree ^56^ visualization of the MST obtained with the INNUENDO-like pipeline is shown. Nodes (i.e. samples) are collapsed at the threshold with highest congruence with CC (508 ADs for this pipeline) and colored according to the ST classification. **B)** Number of partitions obtained by each pipeline at each possible distance threshold. **C)** Clustering stability regions determined for each pipeline. The different blocks of each pipeline start in a different line. Distance thresholds (*x* axis) are presented in log2 scale. **D)** Barplot (top) with the number of samples of the top represented STs (≥ 50 samples) in *L. monocytogenes* dataset, with a swarmplot (bottom) indicating the AD threshold at which each pipeline clusters together all samples of each ST. **E)** Distribution of the AD thresholds at which each pipeline clusters together all samples of a given ST. The outlier STs are indicated above the respective diamond symbol.

#### 2.1 Evaluation of allele-based clustering and comparison of stability regions

Following QC of the initial 3300 *L. monocytogenes* isolates ([Additional file](https://zenodo.org/records/12805751) 1), all pipelines retained >99% of the samples, with the exception of Bionumerics, which only retained ∼95% (Table S1.1 in [Additional file](https://zenodo.org/records/12805751) 2). Despite the intrinsic differences between the pipelines, they generally provided very similar clustering patterns in terms of number of partitions across all possible distance thresholds (Figure 2B). Independently of the schema (Moura ^60^ or Ruppitsch ^61^) and clustering algorithm (GT or HC), the pipelines consistently revealed low stability (i.e., cluster composition considerably changes in proximal distance thresholds) in the highest resolution region (spanning the “outbreak level”), and two main “plateau” regions of high stability (i.e., yielding similar cluster number and composition across a given threshold range), likely reflecting the pathogen population structure and dataset diversity (Figure 2C).

#### 2.2 Evaluation of allele-based clustering congruence with traditional typing

Our results revealed a good correspondence between the first large plateau of stability of all allele-based pipelines and the *L. monocytogenes* ST classification (Figures 2C and 2D), with the highest congruence point being identified between 143 and 190 ADs (Table S1.1 in [Additional file](https://zenodo.org/records/12805751) 2). The maximum CS was very high (∼2.9) but not the maximum possible (3.0), indicating that some of the STs were divided into more than one phylogenetic branch at this level of resolution. Between 75% to 90% (depending on the pipeline) of the 106 STs with two or more samples had all samples grouped in the same cluster, and around half of these were fully isolated, i.e. they were not clustering with another ST. Less than 2% of the STs were split into three or more groups regardless of the pipeline. We then identified the lowest threshold level at which all samples of the same ST cluster together in each pipeline ([Additional file](https://zenodo.org/records/12805751) 3). The majority of the STs congruently cluster below the highest congruence point (albeit at different scales), including prevalent and/or epidemiologically relevant STs, such as ST1, ST5, ST6, ST8 and ST121 (Figure 2D). This in-depth ST-specific analysis also suggested that some STs were consistently polyphyletic regardless of the pipeline, as it is the case of ST7 and ST325 due to the presence of a few same-ST samples (one and four at the highest CS, respectively) clearly clustering apart (Figures 2A and 2E).

Similar results were found when comparing the cgMLST clustering results with *L. monocytogenes* CC typing. In this case, the highest congruence point was identified between 388 and 508 ADs, with the maximum CS (>2.97) being even higher than the one obtained with the ST ([Additional file](https://zenodo.org/records/12805751) 4 and Table S1 in [Additional file](https://zenodo.org/records/12805751) 2). From the 70 CCs with at least two samples, between 60% to 83% (depending on the pipeline) had all samples grouped in the same cluster at the highest congruence point. On average, 89% of these CCs were fully clustered apart, with some of them, including the majority of the CCs with the higher amount of samples, forming a single cluster at thresholds lower than the highest congruence point in all pipelines (Figure S1.1 in [Additional file](https://zenodo.org/records/12805751) 2). In contrast, a few CCs were consistently polyphyletic regardless of the pipeline (Figure S1.1 in [Additional file](https://zenodo.org/records/12805751) 2), although with different signatures. For instance, while CC5 and CC2 had just a few divergent samples leading to the polyphyletic signature, CC4 and CC14 were divided into two main clusters that could only be merged at high threshold levels (considerably above the highest CS) (Figure S1.1 in [Additional file](https://zenodo.org/records/12805751) 2). When comparing the clustering of each CC with the corresponding STs, we noted that CC8 clustered all its samples together at higher AD thresholds than the individual STs (Figure S1.1 in [Additional file](https://zenodo.org/records/12805751) 2), particularly due to ST8 and ST16, which differ from each other by more than 200 ADs but reveal low intra-ST diversity.

#### 2.3 Evaluation of cluster congruence between different pipelines at all threshold levels

Our in-depth pairwise congruence analysis showed a general high concordance between all allele-based pipelines (as exemplified for a pairwise comparison in Figures 3A and 3B, and detailed in Section 2 of [Additional file](https://zenodo.org/records/12805751) 2). Indeed, the AD threshold points with highest concordance (assessed as CS ≥ 2.85) between every two pipelines (“corresponding points”) were observed across all levels of resolution and followed a linear trend (r2 ≥ 0.99) in all comparisons (Figures 3C and 3D and [Additional file](https://zenodo.org/records/12805751) 2, 5 and 6). The slight deviation from a *y=x* scenario (i.e. theoretical situation in which clustering at one level with a pipeline is concordant with the clustering at the exact same level in the other one) revealed differences in their discriminatory power (Figure 3D), which corroborated the need for a fine evaluation at “outbreak” level (Section 2.4).

**Figure 3.**
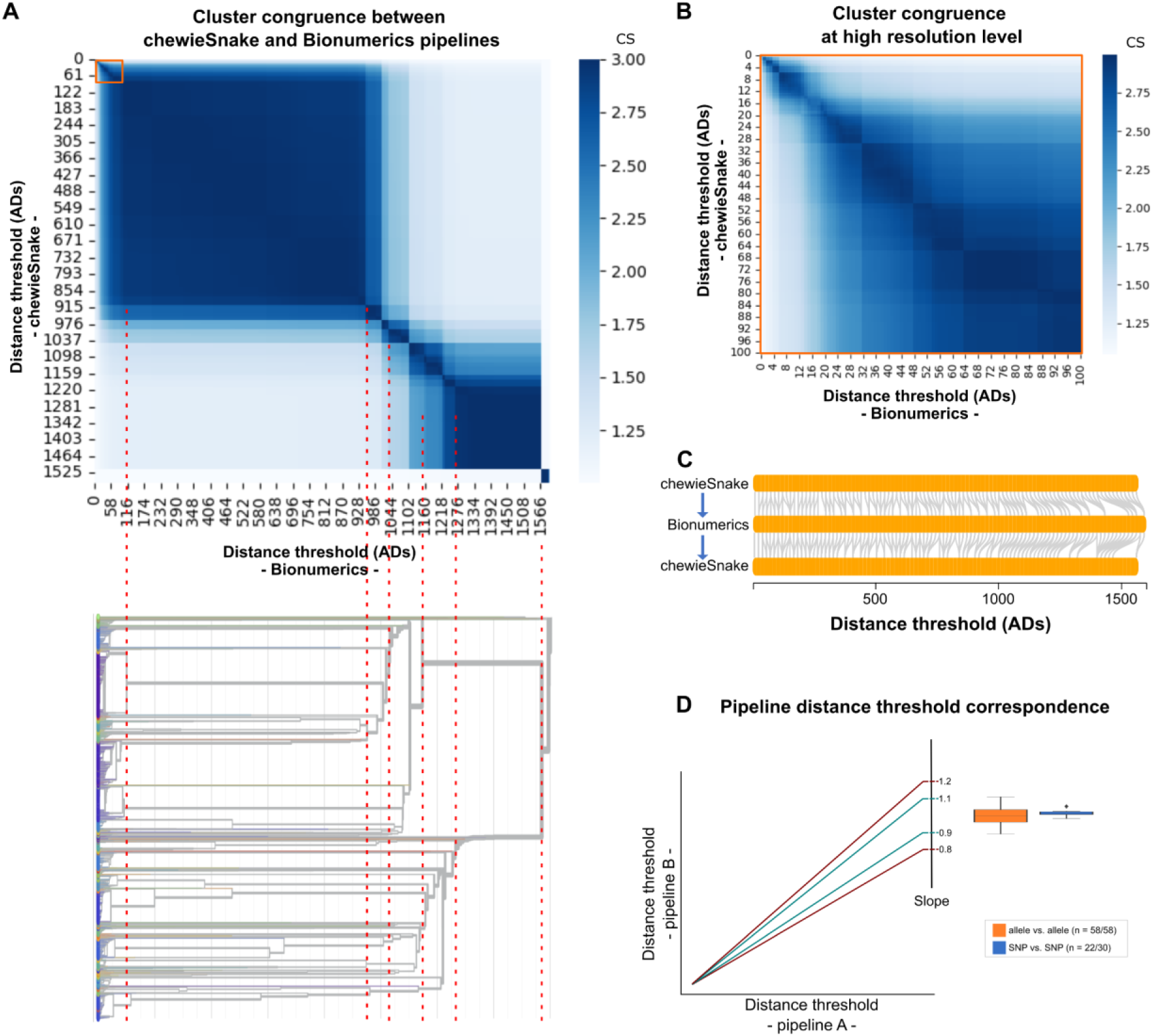
Global evaluation of cluster congruence between different pipelines at all threshold levels for *L. monocytogenes*. **A)** Heatmap with the CS obtained for the comparison of the clustering results obtained at all possible thresholds of two pipelines (details on each pairwise comparison are in [Additional file](https://zenodo.org/records/12805751) 2, with chewieSnake vs. Bionumerics using the HC algorithm being presented here as an example). The orange square highlights the levels of high resolution. To better link the heatmap with the clustering tree, we present an inverted dendrogram (i.e., from the highest to the lowest resolution) showing how the congruence at all levels is related with the phylogenetic structure of the dataset (dendrogram obtained with Bionumerics and visualized in auspice.us ^129^). Dashed red lines connect the CS score and clustering results obtained at similar distance thresholds. **B)** Zoom-in in the high resolution level highlighted in orange in panel A. **C)** Bi-directional correspondence points (gray lines) connecting the thresholds providing similar clustering results (closest threshold with the highest CS; only considered CS ≥ 2.85) by the two pipelines exemplified in panel A. **D)** Representation of the linear trend lines expected for the correspondence points with a theoretical slope deviation of 10% and 20% to be used as scale reference for the boxplots. The slope is used as a proxy to evaluate whether two pipelines yield highly concordant clustering at the exact same level with slopes close to 1 (i.e. *y=x* scenario) reflecting high clustering concordance and similar discriminatory power across all levels. Boxplots present the slope distribution for allele vs. allele (orange) and SNP. vs. SNP (blue) pipeline comparisons for the linear trendlines with a r2 ≥ 0.99, illustrated in [Additional file](https://zenodo.org/records/12805751) 2 and detailed in [Additional file](https://zenodo.org/records/12805751) 6 (“n” refers to the number of comparisons with r2 ≥ 0.99 over the total number of comparisons). The boxplot of the allele vs. SNP scenario is not presented as only 42 out of the 248 comparisons yielded an r2 ≥ 0.99 ([Additional file](https://zenodo.org/records/12805751) 6).

A similar pairwise comparison was performed between SNP-based pipelines for the top-represented STs in the *L. monocytogenes* dataset (ST1, ST5, ST6, ST8 and ST121) with a focus on the clustering obtained at up to a 100 SNPs threshold (detailed in Sections 3 and 4 of [Additional file](https://zenodo.org/records/12805751) 2). We observed an overall concordant clustering regardless of the ST under analysis, as supported by a similar maximum number of possible distance thresholds and number of clusters throughout most levels of resolution ([Additional file](https://zenodo.org/records/12805751) 2). In most comparisons, this high concordance is also illustrated by the nearly symmetric CS matrices with high scores mainly falling within the diagonal (Section 3 of [Additional file](https://zenodo.org/records/12805751) 2 and [Additional file](https://zenodo.org/records/12805751) 6) and by a linear trend of the inter-pipeline corresponding points with a slope very close to 1 (Figure 3D). A notable exception included an intermediate level of resolution with low congruence between CSI Phylogeny and snippySnake/WGSBAC for ST6 and ST8, despite the good concordance at outbreak level (Section 3 in [Additional file](https://zenodo.org/records/12805751) 2).

On the contrary, when comparing allele- and SNP-based pipelines, in most situations, we observed (slightly) asymmetric matrices (see heatmaps in Section 4 of [Additional file](https://zenodo.org/records/12805751) 2), with similar clustering (assessed as high CS) often obtained at higher SNP threshold levels than ADs. These results indicate that SNP-based pipelines, when using the same ST reference for read mapping (a strategy in place in some laboratories), tend to provide a higher discriminatory power than cgMLST pipelines, even though this might not be applicable for all STs. For instance, this asymmetric trend was not so evident for STs 6 and 8 (Section 4 in [Additional file](https://zenodo.org/records/12805751) 2), as similar cluster composition was observed at similar SNP and AD threshold levels. In general, we found a low number of corresponding points in the pairwise comparisons (assessed at up to 100 ADs/SNPs), which rarely (30/246) yielded a linear trend (i.e. with r2 ≥ 0.99, [Additional file](https://zenodo.org/records/12805751) 6), thus challenging the overall comparison of the discriminatory power through this approach (Figure 3D). Still, there was a high concordance at outbreak level in most situations, as seen in the heatmaps (Section 4 in [Additional file](https://zenodo.org/records/12805751) 2), showing that a more detailed analysis for outbreak detection is more informative about pipeline performance when comparing allele- and SNP-based pipelines (see section 3.4).

As a complementary exercise, a SNP-based pipeline (“CSI Phylogeny” ^62^) was also applied in a dataset combining the five STs assessed individually using the reference of ST6. As expected, the discriminatory power dropped, reaching a level even lower than the one provided by allele-based pipelines (Sections 3 and 4 of [Additional file](https://zenodo.org/records/12805751) 2). This points out that read mapping against a single reference for multiple STs does not provide enough resolution for routine surveillance and outbreak investigation.

#### 2.4 Concordance for outbreak detection

Allele-based approaches are the most commonly applied for *L. monocytogenes* outbreak detection, and the distance threshold corresponding to 7 ADs is conventionally used to determine potential outbreak-related samples ^60,63^. As such, we used this threshold to identify the potential outbreak-related clusters determined by each allele-based pipeline for the *L. monocytogenes* dataset. Each pipeline detected between 310 to 340 clusters at 7 ADs, from which ∼94.2% had similar composition in at least two pipelines and 5.8% were exclusively detected by a single pipeline ([Additional file](https://zenodo.org/records/12805751) 7). Only ∼50% of the clusters detected by a given pipeline was also detected with the exact same composition in all pipelines, but this value is highly influenced by the diversity of the studied pipelines and by the use of a static cut-off. For instance, this value would increase to ∼72%, if the most discrepant pipeline was removed (MentaLiST), and to almost 90%, if only same-schema pipelines were compared ([Additional file](https://zenodo.org/records/12805751) 7).

As these results are impacted by the use of a static threshold, we identified the minimum threshold (ADs or SNPs) at which each 7 AD cluster would be detected by the other pipelines (see the Methods section for details) ([Additional file](https://zenodo.org/records/12805751) 8). This analysis yielded 316 clusters that, once detected at 7 ADs by at least one pipeline, were detected by all pipelines regardless of the threshold ([Additional file](https://zenodo.org/records/12805751) 8). As expected, most of these clusters were detected at ≤ 7 ADs in all pipelines, or at higher threshold levels close to 7 ADs (Figure 4A). The difference between the AD thresholds required by the different allele-based pipelines to detect each “outbreak-level” cluster had a median of 2 ADs (1 AD, if MentaLiST is excluded), with a minimum of 0 and maximum of 24 ADs (Figure 4B). Without MentaLiST, the pairwise comparisons of the remaining pipelines showed that the overlap of clusters detected at 7 ADs with the exact same composition was 84.5%, on average, a value that increased to 93.0%, when applying a flexible threshold of up to 2 ADs above (Figures 4C and 4D and [Additional file](https://zenodo.org/records/12805751) 9). The cluster congruence at this level of resolution is influenced by the cgMLST schema used, with pipelines using the same schema yielding more similar results (Figure 4C and 4D). This is showcased through the analysis of the threshold flexibilization (Figure 4B), in which the overlap increases to 95.2% and 97.5% when only Moura or Ruppitsch pipelines are compared (Figure 4B), respectively. In a case scenario where the recommended static thresholds for each schema would be applied, i.e. 7 ADs for Moura ^60^ and 10 ADs for Ruppitsch ^61^, the overlap of “outbreak signals’’ between pipelines running different schemas would be considerably lower than the one obtained with a flexible approach. We also tested the application of a more stringent threshold (4 ADs) to identify isolates with more compelling evidence of being part of the same outbreak, followed by the application of a higher cut-off (7 ADs) for identifying probable cases, as previously proposed ^63^. This exercise showed that clusters defined at 4 ADs by a given pipeline are very often captured with the same composition by any other pipeline with a threshold of up to 7 ADs, with the exception of MentaLiST ([Additional file](https://zenodo.org/records/12805751) 9).

**Figure 4.**
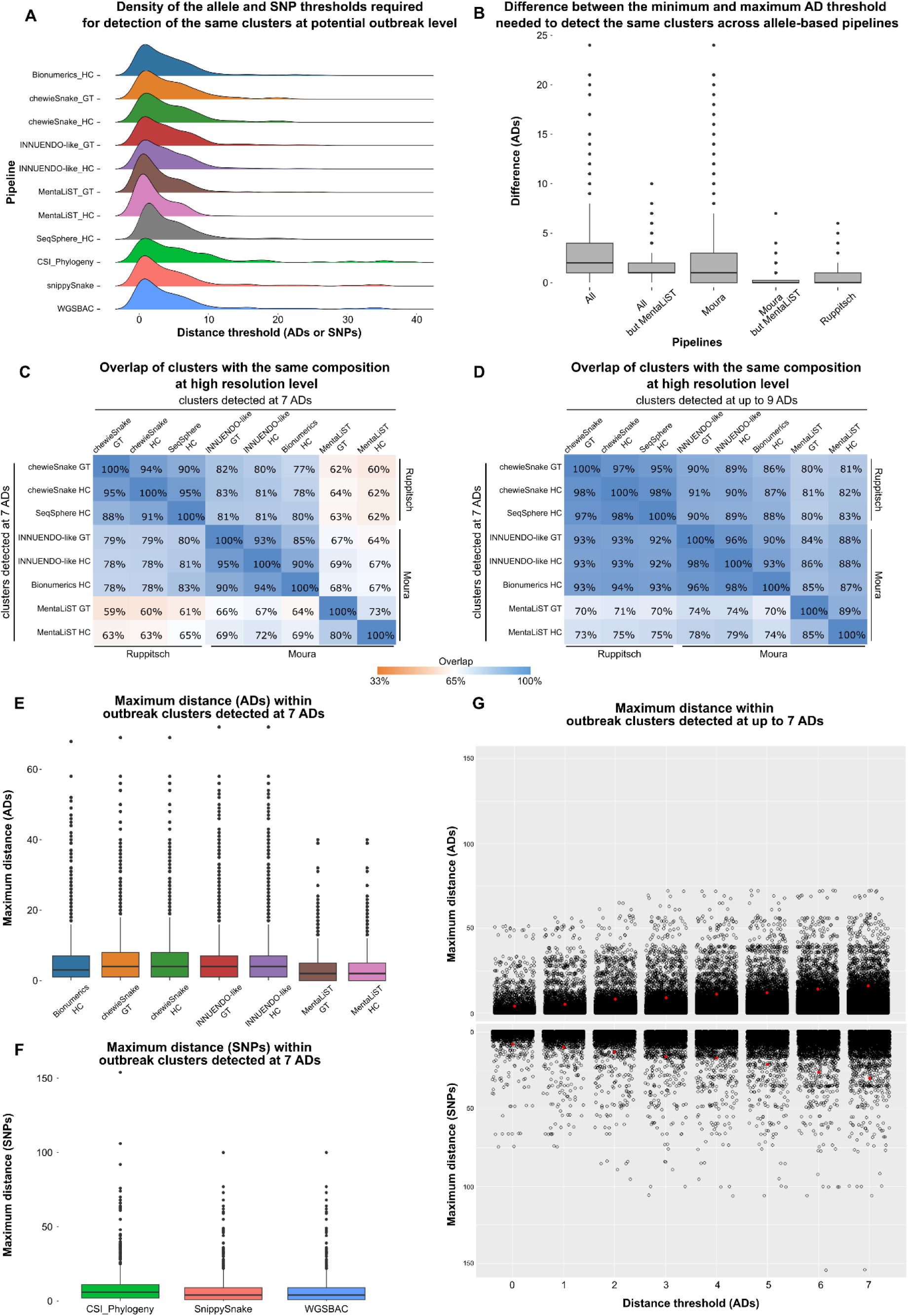
Evaluation of the performance of the genome-scale pipelines in the identification of potential outbreak-related isolates for the *L. monocytogenes* dataset. **A)** Density of the distance thresholds (ADs for allele-based pipelines and SNPs for SNP-based pipelines) required for the identification of clusters detected by at least one allele-based pipeline at 7 ADs. Only clusters having the same composition in all allele-based pipelines were included (n = 316). The results of SNP-based pipelines only include the clusters corresponding to the top 5 STs and were obtained using an ST-specific reference. **B)** Distribution of the difference between the minimum and maximum AD threshold needed to detect the same clusters across allele-based pipelines, using the same clusters as described in panel A. **C)** Overlap between the genetic clusters detected by allele-based pipelines at 7 ADs. **D)** Overlap between the genetic clusters detected by one pipeline at 7 ADs and those detected by the others at ≤ 9 ADs. **E)** Distribution of the maximum ADs within each “outbreak-level” cluster identified at 7 ADs by the respective pipeline. **F)** Distribution of the maximum SNP distances within “outbreak-level” cgMLST clusters identified by at least an allele-based pipeline at 7 ADs. **G)** Maximum allele (top) or SNP (bottom) distance threshold within each cluster determined by at least an allele-based pipeline at up to 7 ADs. The red dot indicates, for each threshold, the 95^th^ percentile.

When looking at the genomic diversity (SNPs/ADs) within the cgMLST “outbreak clusters” (7 ADs), our results showed that the maximum allele/SNP distances increase with the size of the cluster and are larger when looking at SNPs (Figure 4E and 4F). These results are consistent with the previous observation that higher SNP thresholds (which increase alongside with AD thresholds) are needed to identify cgMLST clusters with the exact same composition (Figure 4G), while suggesting that, in general, SNP-based pipelines run per ST leverage good resolution to discriminate strains from the same outbreak.

### 3. Salmonella enterica

*Salmonella enterica* dataset (Figure 5A) was analyzed with seven allele-based and four SNP-based pipelines (Table 1 and Tables S1.1 and S1.2 in Additional file 10).

**Figure 5.**
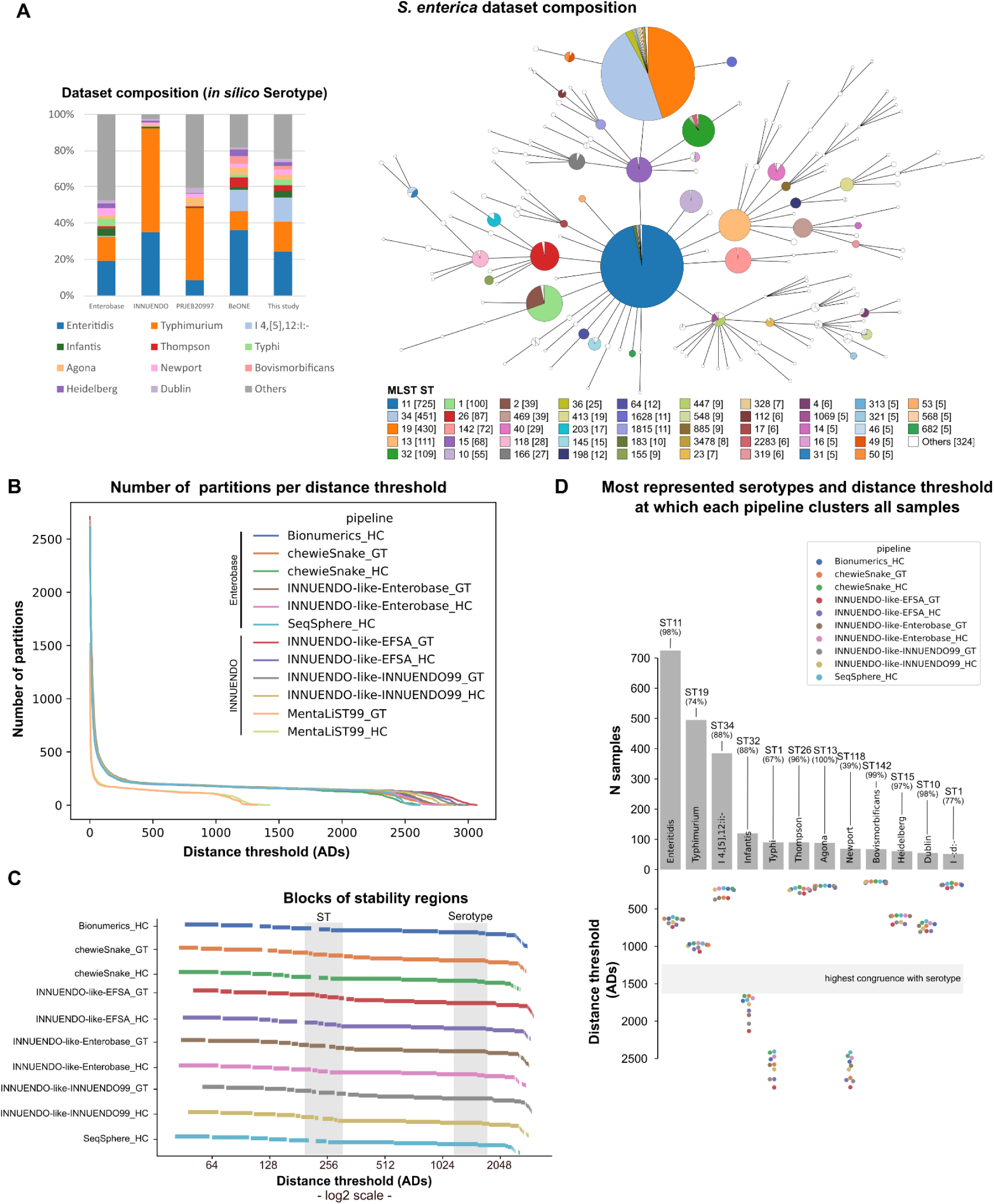
Assessment of allele-based clustering at all possible threshold levels for *S. enterica* and congruence with traditional serotype. **A)** Composition of the *S. enterica* dataset used in this study in terms of serotype and in comparison with the composition of the datasets of previous studies (INNUENDO ^20^ and BioProject PRJEB20997 ^130^), and the Enterobase database as of November 2021 ^64^. A GrapeTree ^56^ visualization of the MST obtained with the INNUENDO-like-INNUENDO99 pipeline is shown. Nodes (i.e. samples) are collapsed at the threshold with highest congruence with serotype (1514 ADs for this pipeline) and colored according to the ST classification. **B)** Number of partitions obtained by each pipeline at each possible distance threshold. **C)** Clustering stability regions determined for each pipeline. The different blocks of each pipeline start in a different line. Distance thresholds (*x* axis) are presented in log2 scale. **D)** Barplot (top) with the number of samples of the top represented serotypes (≥ 50 samples) in *S. enterica* dataset, with a swarmplot (bottom) indicating the AD threshold at which each pipeline clusters together all samples of each serotype.

#### 3.1 Evaluation of allele-based clustering and comparison of stability regions

Following QC of the initial 2974 *S. enterica* isolates ([Additional file](https://zenodo.org/records/12805751) 1), a maximum of 2% of the isolates were filtered out by each pipeline (Table S1.1 in [Additional file](https://zenodo.org/records/12805751) 10). Despite the intrinsic differences between the pipelines, they generally provided very similar clustering patterns in terms of number of clusters across all possible partitions, with the exception of MentaLiST (Figure 5B). Given the outlier behavior of MentaLiST that, as seen for *L. monocytogenes*, has a considerable negative impact in pipeline comparisons and interpretation of the results, we decided to remove this tool from all downstream analyses, including for *E. coli* and *C. jejuni*.

Independently of the schema (Enterobase ^64^ or INNUENDO ^20^) and clustering algorithm (GT or HC), the pipelines consistently revealed low stability in the region of high resolution spanning the “outbreak level”, but several regions of high stability could be identified with good correspondence between pipelines, likely reflecting the pathogen population structure and dataset diversity (Figure 5C). The largest stability region (covering ∼690 subsequent AD thresholds) was similar between pipelines, occurred between ∼1000 and ∼1700 ADs and corresponded to the largest stability region detected in a previous study ^65^.

#### 3.2 Assessment of allele-based clustering congruence with traditional typing

Our results revealed a good correspondence between the largest stability region detected in all allele-based pipelines and *S. enterica* serotype classification (Figures 5C and 5D), with the highest congruence point being identified between 1261 and 1663 ADs (CS ∼2.3) (Table S1.1 in [Additional file](https://zenodo.org/records/12805751) 10). From the 91 serotypes with at least two samples, between 44% to 68% are grouped in the same cluster at the highest congruence point. This observation is aligned with the results of a large study in which 70.1% of the analyzed serotypes mapped to a single cgMLST cluster in an equivalent stability region ^65^. Still, when focusing on those serotypes having a one-to-one cluster correspondence (i.e. the whole cluster corresponds to all samples of a serotype) in our study, this number decreased to about 30% of the serotypes, regardless of the pipeline. Remarkably, the one-to-one correspondence was detected for the majority of the most prevalent serotypes, although the lowest threshold needed to collapse all samples was quite diverse across serotypes (Figure 5D). Our analysis also revealed that between 8% to 25% of all serotypes are split into three or more clusters, suggesting the existence of polyphyletic serotypes. Among these, we highlight the Thompson and Newport serotypes ([Additional file](https://zenodo.org/records/12805751) 11), for which a threshold of more than 2400 ADs was required to collapse all the respective samples, which is in accordance with their previously reported multi-lineage nature ^65–68^.

Regarding the congruence with the ST, the highest congruence point was identified between 205 and 310 ADs (CS ∼2.6), always falling within a pipeline stability region (Figure 5C, Table S1.1 in [Additional file](https://zenodo.org/records/12805751) 10). From the 112 STs with more than two samples, depending on the pipeline, between 73% to 85% (82 to 95 STs) were grouped in a single cluster at the highest congruence point. On average, 59% of the STs exactly corresponded to a single cluster and a small proportion (between 4% and 9%) were split into three or more clusters. When looking at the earliest threshold to merge all samples of a given ST, some STs clustered considerably below the highest CS (e.g., ST34 and ST26) while others revealed high intra-ST heterogeneity (e.g., ST11 and ST15) ([Additional file](https://zenodo.org/records/12805751) 12 and Figure S1.1 in [Additional file](https://zenodo.org/records/12805751) 10).

In this dataset, Enteritidis serotype is mainly composed of ST11 samples, and our results revealed a good concordance between the thresholds required to merge either Enteritidis or ST11 samples, with ST11 requiring a slightly higher threshold due to few samples not predicted as Enteritidis (Figure 5D and Figure S1.1 in [Additional file](https://zenodo.org/records/12805751) 10). Regarding the samples classified *in sílico* as Typhimurium, they were segregated into three main STs (ST19, ST34 and ST36) with different levels of intra-ST diversity. This likely justifies why all Typhimurium samples were only collapsed in a single cluster at a high threshold (Figure 5D and Figure S1.1 in [Additional file](https://zenodo.org/records/12805751) 10). Still, we cannot discard that this value is overestimated due to ST34 samples that were classified as Typhimurium instead of its most common classification within the Typhimurium monophasic variant 4,[5],12:i:- (here treated as an independent serogroup). Finally, Infantis serotype has a high diversity and a potential polyphyletic signature, which contrasts with its dominant ST (ST32). This is due to a few non-ST32 Infantis samples present in this dataset (Figure 5D and Figure S1.1 in [Additional file](https://zenodo.org/records/12805751) 10).

#### 3.3 Evaluation of cluster congruence between different pipelines at all threshold levels

Our in-depth pairwise congruence analysis showed a general high concordance between all allele-based pipelines (as exemplified for a pairwise comparison in Figures 6A and 6B, and detailed in Section 2 of [Additional file](https://zenodo.org/records/12805751) 10). Indeed, the AD threshold points with highest concordance (assumed as CS ≥ 2.85) between every two pipelines (“corresponding points”) were observed across all levels of resolution and followed a linear trend (r2 >= 0.99) in all comparisons (Figure 6C and 6D, [Additional file](https://zenodo.org/records/12805751) 10, 13 and 14). Despite the good inter-pipeline concordance (even at low threshold levels), differences in the discriminatory power were still observed, as shown by deviations from a *y=x* scenario (Figure 6D). A fine-tuned analysis about pipeline performance and comparability at “outbreak” level is presented below (section 3.4).

**Figure 6.**
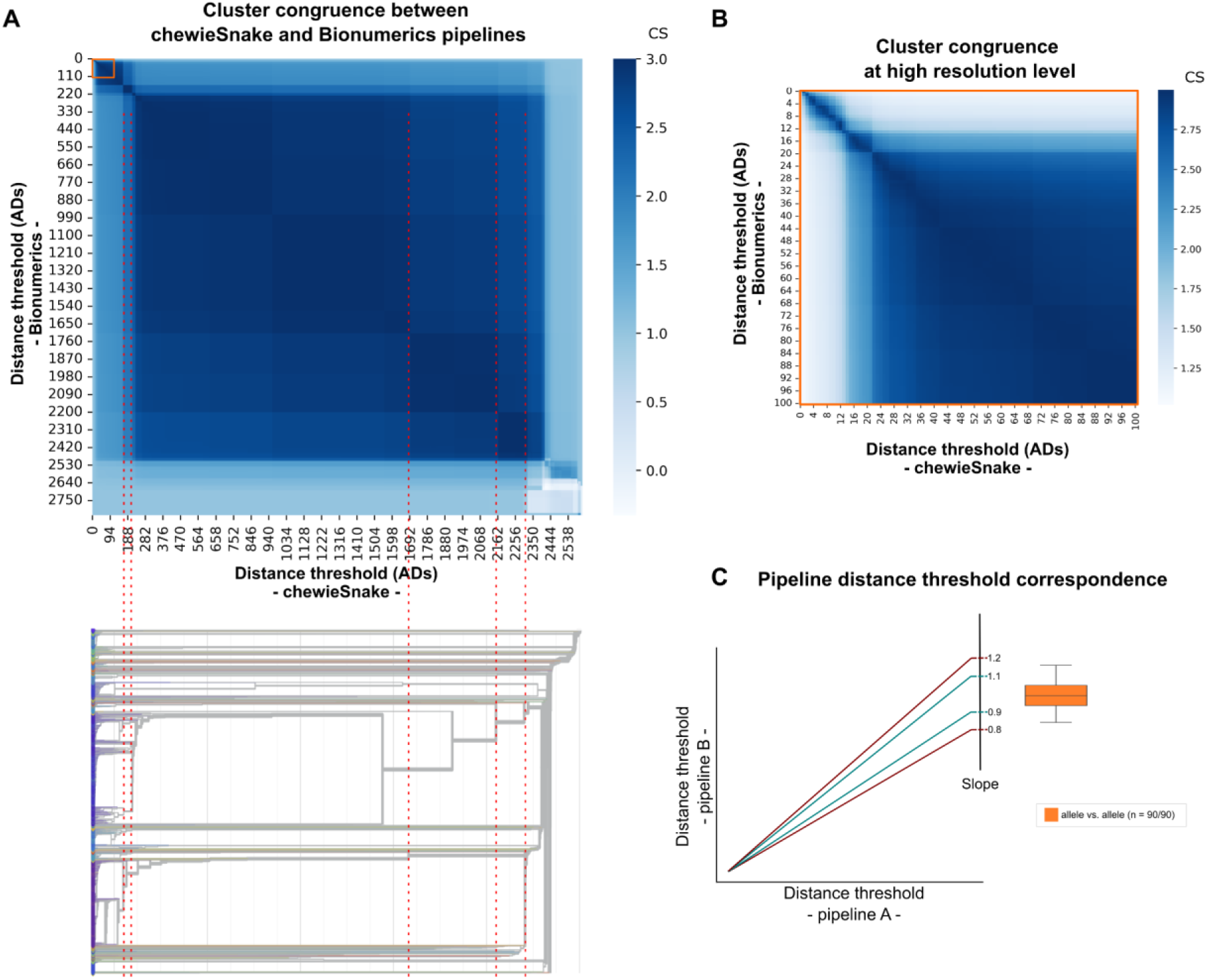
Global evaluation of cluster congruence between different pipelines at all threshold levels for *S. enterica* dataset. **A)** Heatmap with the CS obtained for the comparison of the clustering results obtained at all possible thresholds of two pipelines (details on each pairwise comparison are in [Additional file](https://zenodo.org/records/12805751) 10, with Bionumerics vs. chewieSnake using the HC algorithm being presented here as an example). The orange square highlights the levels of high resolution. To better link the heatmap with the clustering tree, we present an inverted dendrogram (i.e., from the highest to the lowest resolution) showing how the congruence at all levels is related with the phylogenetic structure of the dataset (dendrogram obtained with chewieSnake and visualized in auspice.us ^129^). Dashed red lines connect the CS score and clustering results obtained at similar distance thresholds. **B)** Zoom-in in the high resolution level highlighted in orange in panel A. **C)** Representation of the linear trend lines expected for the correspondence points with a theoretical slope deviation of 10% and 20% to be used as scale reference for the boxplots. The slope is used as a proxy to evaluate whether two pipelines yield highly concordant clustering at the exact same level with slopes close to 1 (i.e. *y=x* scenario) reflecting high clustering concordance and similar discriminatory power across all levels. The boxplot presents the slope distribution for allele vs. allele (orange) pipeline comparisons for the linear trendlines with a r2 ≥ 0.99, illustrated in [Additional file](https://zenodo.org/records/12805751) 10 and detailed in [Additional file](https://zenodo.org/records/12805751) 14 (“n” refers to the number of comparisons with r2 ≥ 0.99 over the total number of comparisons). The boxplots of the SNP vs. SNP and allele vs. SNP scenarios are not presented as only 4/14 and 6/160 comparisons yielded an r2 ≥ 0.99 ([Additional file](https://zenodo.org/records/12805751) 14).

Regarding the SNP-based pipelines, the analysis was conducted with a focus on the clustering obtained at up to a 100 SNPs threshold for the top-represented serotypes, namely Enteritidis, Typhimurium and Infantis, in all pipelines, except for SnapperDB, as the partner institute could only run it for Typhimurium and Infantis subdatasets. As SnippySnake and WGSBAC yielded matching partitions at all levels, only SnippySnake results are presented. SNP-based pipelines revealed considerable differences in the maximum number of thresholds (detailed in Sections 3 and 4 of [Additional file](https://zenodo.org/records/12805751) 10), which are more pronounced between SnapperDB and the other pipelines. SnapperDB excluded samples that, although being *in sílico* predicted as Typhimurium or Infantis, were phylogenetically distant from the remaining ones, reducing the number of informative sites in the core alignment and, consequently, leading to a lower maximum number of partitions for these serotypes in this pipeline but a higher resolution power. This variability in sample inclusion/exclusion challenged the assessment of the congruence at all levels between pipelines, as illustrated by the asymmetry of the heatmaps (Section 3 in [Additional file](https://zenodo.org/records/12805751) 10). Still, most pairwise comparisons were informative, as revealed by the often observation of concordant clustering results, especially at lower threshold levels.

When comparing allele- and SNP-based pipelines, we observed (slightly) asymmetric matrices (see heatmaps in Section 4 of [Additional file](https://zenodo.org/records/12805751) 10), often deviating towards high SNP thresholds (i.e. high CSs are observed when the SNP threshold is higher than the corresponding AD threshold) (e.g. Figure S4.1.6 in Section 4 of [Additional file](https://zenodo.org/records/12805751) 10). As such, the SNP-based pipelines, when using the same serotype reference for read mapping, tend to provide a higher discriminatory power than cg/wgMLST pipelines, even though this might not be applicable for all serotypes and pipelines. For instance, this trend was inverted for Enteritidis serotype when using SnippySnake pipeline (e.g. Figure S4.3.2 in Section 4 of [Additional file](https://zenodo.org/records/12805751) 10). In general, we found a low number of corresponding points in the pairwise comparisons (assessed at up to 100 ADs/SNPs) and the few identified points did not follow a linear trend (i.e. with r2 ≥ 0.99, [Additional file](https://zenodo.org/records/12805751) 14), thus hampering a broad assessment of the discriminatory power. Still, the observed concordance trends at outbreak level showed that a more detailed analysis for outbreak detection is more informative about pipeline performance when comparing allele- and SNP-based pipelines (as addressed in section 3.4).

#### 3.4 Concordance for outbreak detection

Allele-based approaches are the most commonly applied for *S. enterica* outbreak detection, but the method and distance threshold used to determine a possible outbreak-related cluster usually varies between laboratories and the inclusion criteria for outbreak is usually set during investigation. The INNUENDO project proposed a dynamic threshold of 0.43% of the cgMLST schema, corresponding to 14 ADs in the INNUENDO cgMLST schema, due to its good concordance with clusters of epidemiologically verified isolates ^20^. We used this 0.43% threshold (which translates into 14 ADs in all pipelines) to start exploring the pipeline congruence at potential outbreak level.

Each pipeline detected between 216 to 254 clusters at 14 ADs, from which, on average, 95.9% had similar composition in at least two pipelines and 4.1% were exclusively detected by a single pipeline ([Additional file](https://zenodo.org/records/12805751) 15). On average, 62.6% of the clusters detected by a given pipeline was also detected with the exact same composition by all remaining pipelines. However, this value is highly influenced by the diversity of the studied pipelines and by the use of a static cut-off. Indeed, this value would increase to almost 75% if only same-schema pipelines are compared ([Additional file](https://zenodo.org/records/12805751) 15). We further evaluated the minimum threshold level (ADs or SNPs) at which each 14 AD cluster would be detected by the other pipelines. This analysis yielded a total of 255 clusters that, once detected at 14 ADs by at least one pipeline, were detected by all pipelines regardless of the threshold ([Additional file](https://zenodo.org/records/12805751) 16). As expected, most of these clusters were detected at a threshold ≤ 14 ADs in all pipelines, or at higher threshold levels close to 14 ADs. At SNP level, two different profiles were observed (Figure 7A). While SnippySnake showed a density profile similar to allele-based pipelines (i.e. a threshold of 14 SNPs would be enough to capture most of the clusters determined at 14 ADs), the other two SNP-based pipelines (SnapperDB and CSI Phylogeny) very often required higher SNP thresholds to merge isolates belonging to the same cluster (Figure 7A). The difference between the AD thresholds required by the different allele-based pipelines to detect each “outbreak-level” cluster had a median of 2 ADs, with a minimum of 0 and maximum of 14 ADs. This trend is less influenced by the clustering algorithm rather than the cg/wgMLST schema, as a median of only 1 AD difference is observed when comparing same-schema pipelines (Figure 7B). Looking at pairwise comparisons between all pipelines, our results showed that the overlap of clusters detected at 14 ADs with the exact same composition was 79.0%, on average, a value that increased to 89.8% when applying a flexible threshold of up to 2 ADs above (Figures 7C and 7D, [Additional file](https://zenodo.org/records/12805751) 17). Importantly, the overall pairwise congruence only slightly decreased when testing thresholds with higher resolution, namely 85.0% for ≤ 10 ADs, as commonly defined ^31,69^, or 81.6% for ≤ 5 ADs, a strict threshold that has recently been used for case definition in multicountry outbreaks ^70,71^ ([Additional file](https://zenodo.org/records/12805751) 17).

**Figure 7.**
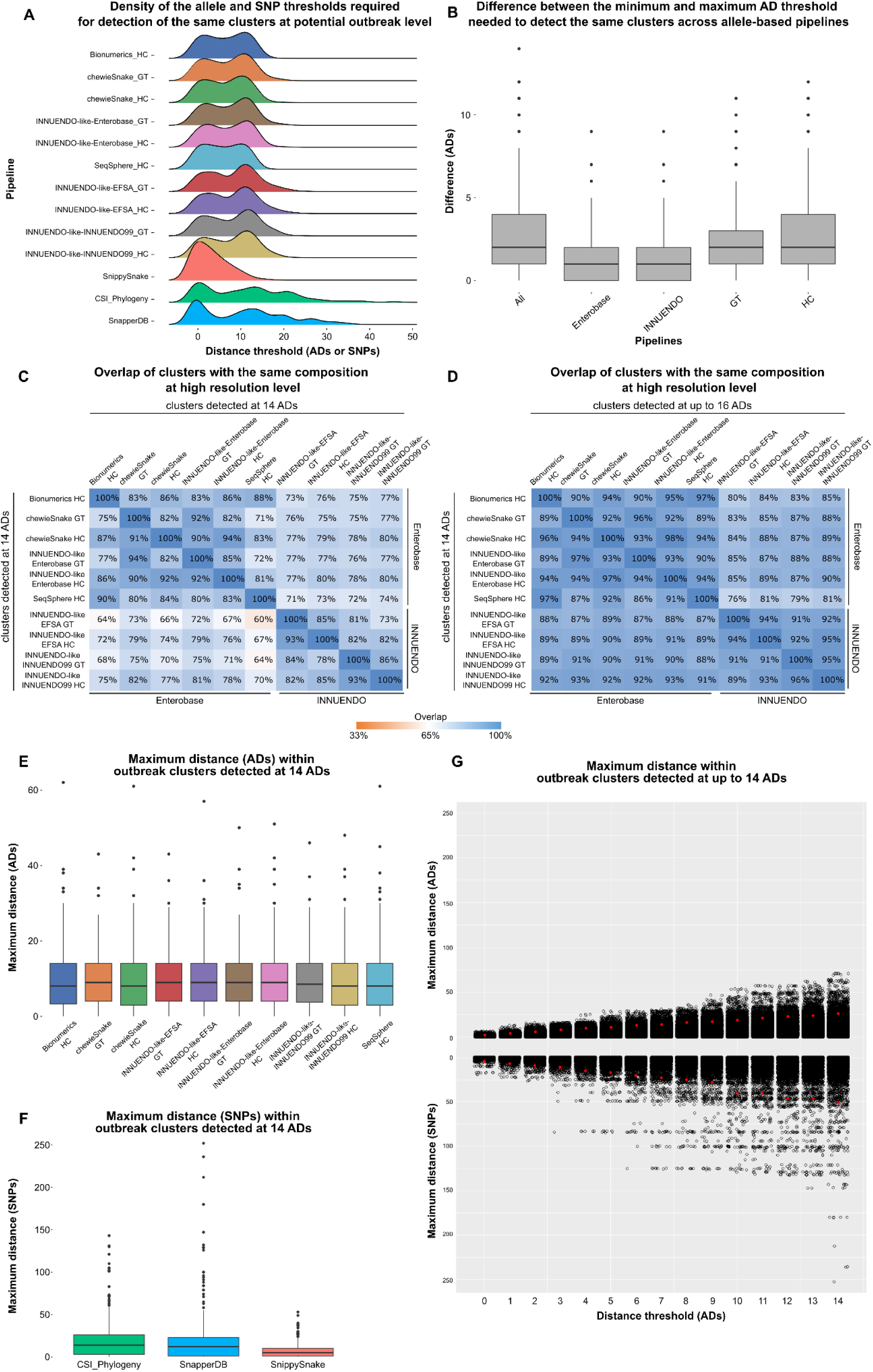
Evaluation of the performance of the genome-scale pipelines in the identification of potential outbreak-related isolates for the *S. enterica* dataset. **A)** Density of the distance thresholds (ADs for allele-based pipelines and SNPs for SNP-based pipelines) required for the identification of clusters detected by at least one allele-based pipeline at 14 ADs. Only clusters having the same composition in all allele-based pipelines were included (n = 255). The results of SNP-based pipelines only include the clusters corresponding to the top 3 serotypes and were obtained using a serotype-specific reference. **B)** Distribution of the difference between the minimum and maximum AD threshold needed to detect the same clusters across allele-based pipelines, using the same clusters as described in panel A. **C)** Overlap between the genetic clusters detected by allele-based pipelines at 14 ADs. **D)** Overlap between the genetic clusters detected by one pipeline at 14 ADs and those detected by the others at ≤ 16 ADs. **E)** Distribution of the maximum ADs within each cluster identified at 14 ADs by the respective pipeline. **F)** Distribution of the maximum SNP distances within cgMLST clusters identified by at least an allele-based pipeline at 14 ADs. **G)** Maximum allele (top) or SNP (bottom) distance threshold within each cluster determined by at least an allele-based pipeline at up to 14 ADs. The red dot indicates, for each threshold, the 95^th^ percentile.

When looking at the genomic diversity (SNPs/ADs) within the cg/wgMLST clusters at 14 ADs, our results showed that allele-based pipelines behave similarly, with the maximum intra-cluster distance increasing with the size of the cluster, as expected (Figure 7E). Although this trend is also seen at SNP level, SnapperDB and CSI Phylogeny required higher SNP thresholds than SnippySnake to identify cgMLST clusters with the exact same composition (Figure 7A), and yielded a higher SNP diversity within the clusters (Figure 7F). The evaluation of intra-cluster diversity across incremental distance thresholds also shows that SNP-based pipelines capture a higher diversity than allele-based pipelines. For example, 95% of the clusters detected at 5 ADs by at least one allele-based pipeline were composed by strains that diverge by no more than 11 alleles, a value that increases to 17 when assessed in terms of SNPs (Figure 7G).

Finally, we conducted an additional exercise with the pipeline running an wgMLST schema (INNUENDO-like-INNUENDO99) to explore the potential gain in resolution to discriminate potential outbreak isolates (as assessed by cgMLST) when increasing the number of wgMLST loci under comparison, aligned with a previously explored rationale ^20,55^. Regardless of the clustering algorithm, this approach resulted in an average increase of 6 ADs in the maximum pairwise distances observed between the isolates of the same original cgMLST cluster ([Additional file](https://zenodo.org/records/12805751) 18), demonstrating the clear increase in resolution provided by the dynamic extension of the cgMLST schema with wgMLST loci shared by the same-outbreak isolates.

### 4. Escherichia coli

*Escherichia coli* dataset (Figure 8A) was analyzed with seven allele-based and two SNP-based pipelines (Table 1 and Tables S1.1 and S1.2 in Additional file 19).

**Figure 8.**
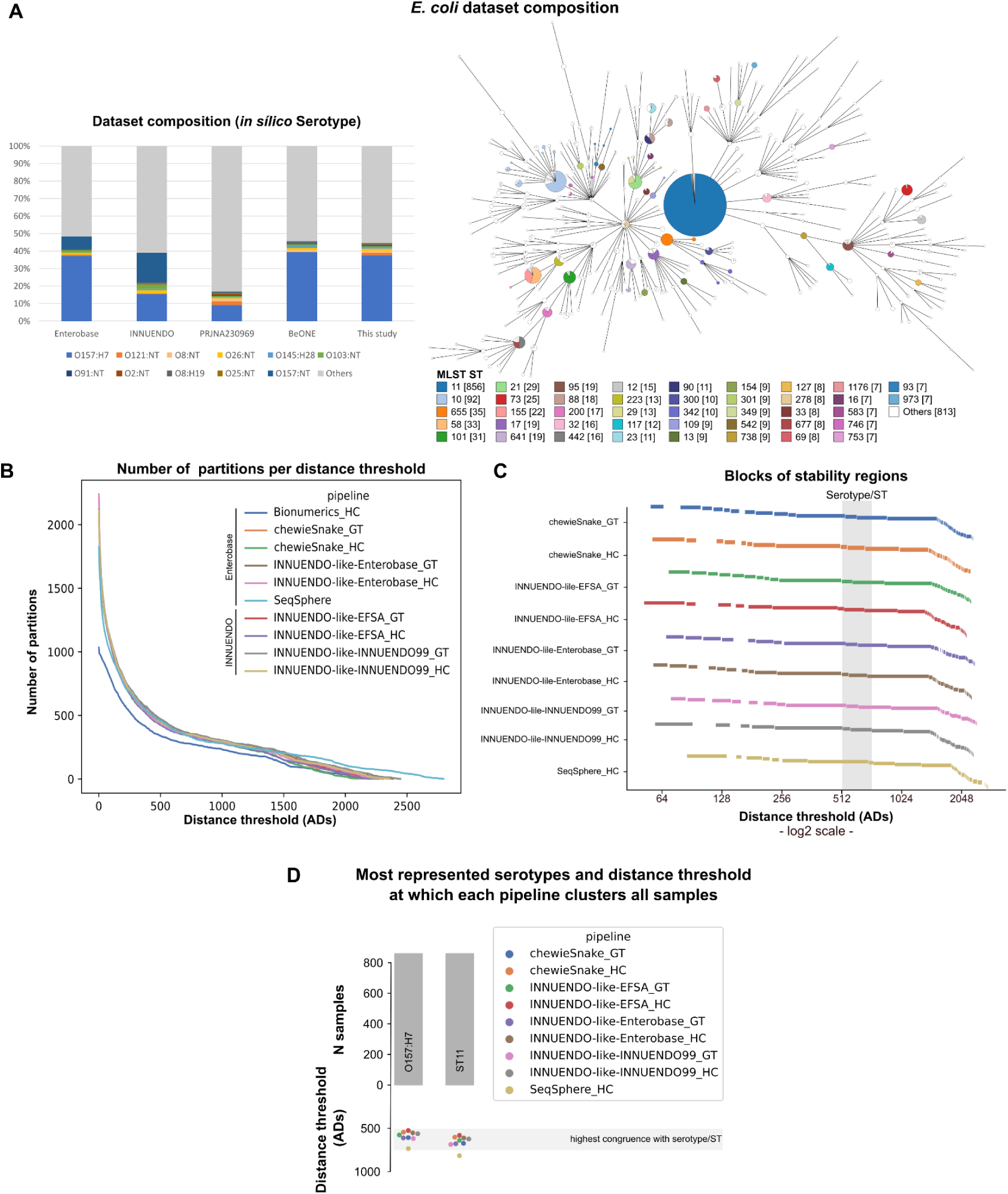
Assessment of allele-based clustering at all possible threshold levels for *E. coli* and congruence with traditional serotype. **A)** Composition of the *E. coli* dataset used in this study in terms of serotype and in comparison with the composition of the datasets of previous studies (INNUENDO ^20^ and BioProject PRJNA230969 ^131,132^), and the Enterobase database as of November 2021 ^64^. A GrapeTree ^56^ visualization of the MST obtained with the INNUENDO-like-INNUENDO99 pipeline is shown. Nodes (i.e. samples) are collapsed at the threshold with highest congruence with serotype (620 ADs for this pipeline) and colored according to the ST classification. **B)** Number of partitions obtained by each pipeline at each possible distance threshold. **C)** Clustering stability regions determined for each pipeline. The different blocks of each pipeline start in a different line. Distance thresholds (*x* axis) are presented in log2 scale. **D)** Barplot (top) with the number of samples of the most represented serotype (O157:H7) and ST (ST11) in *E. coli* dataset, with a swarmplot (bottom) indicating the AD threshold at which each pipeline clusters together all samples of each of them.

#### 4.1 Evaluation of allele-based clustering and comparison of stability regions

Following QC of the initial 2307 *E. coli* isolates ([Additional file](https://zenodo.org/records/12805751) 1), all pipelines retained more than 99% of the samples, with the exception of SeqSphere and Bionumerics, which only retained 89.99% and 46.25%, respectively (Table S1.1 in [Additional file](https://zenodo.org/records/12805751) 19). As all pipelines used an inclusion criteria of at least 95% cgMLST loci called, this result was most likely linked to the allele caller than to the schema. Indeed, all pipelines relying on chewBBACA ^72^ retained >99% of the samples, even when using the same schema as SeqSphere and Bionumerics (the Enterobase schema ^64^). Given the very low number of samples that passed the QC of Bionumerics, this pipeline was excluded from the analysis of the *E. coli* dataset. Despite the intrinsic differences between the remaining pipelines, they generally provided very similar clustering patterns in terms of number of clusters across all possible thresholds, with the exception of SeqSphere (Figure 8B), which presented a deviating pattern possibly due to the removal of ∼10% of the samples.

Independently of the schema (Enterobase ^64^ or INNUENDO ^20^) and clustering algorithm (GT or HC), the pipelines consistently revealed low stability in the region of high resolution spanning the “outbreak level”. Beyond this region, multiple regions of high stability could be identified across all levels, including a first high resolution region (around 60 to 120 ADs), likely reflecting the dataset diversity, as discussed below (Figure 8C).

#### 4.2 Evaluation of allele-based clustering congruence with traditional typing and WGS-derived pathogen main lineages

This assessment is highly influenced by the often incompleteness in the inference of O and H antigens and, specially, by the dominance of serotype O157:H7 strains (almost all from ST11) in the dataset, which reflects the bias towards this pathogenic *E. coli* in public databases (Figure 8A). As consequence, for all pipelines, the highest congruence point was the same for serotype (CS ∼2.7) and ST (CS ∼2.9) (Table S1.1 in [Additional file](https://zenodo.org/records/12805751) 19) and corresponded to the minimum threshold needed to collapse all O157:H7 and ST11 strains, ranging between 545 and 738 ADs (Figure 8D). This point revealed a good correspondence with one of the largest stability regions (Figures 8C and 8D), but, due to the *E. coli* diversity bias, this result should be eyed with caution, if intended to inform nomenclature design.

Regarding the less abundant serotypes, we observed very different profiles, with those needing a higher threshold to collapse all samples being also the ones with the highest ST diversity. For instance, among the ones with ≥10 isolates, O145:H28 (including ST32 and ST137) was collapsed at around 350 ADs, while O8:H19 (including ST88, ST90, ST201 and ST3233) required around 2250 ADs to merge all same-serotype isolates (Figure 8D, [Additional files](https://zenodo.org/records/12805751) 20 and 21). Also, the lack of one-to-one correspondence was also illustrated by the detection of some STs (e.g. ST88 and ST90) comprising isolates from different serotypes.

Finally, we assessed the congruence between cg/wgMLST clustering and WGS-derived pathogen main lineages inferred through PopPUNK ^73^. For this *E. coli* dataset, PopPUNK clustering had a high congruence (CS >2.97) at allelic distance thresholds (723 to 1002 ADs) above the level with highest congruence with serotype and ST (Table S1.1 in [Additional file](https://zenodo.org/records/12805751) 19).

#### 4.3 Evaluation of cluster congruence between different pipelines at all threshold levels

Our in-depth pairwise congruence analysis showed a high concordance between all allele-based pipelines (as exemplified for a pairwise comparison in Figures 9A and 9B, and detailed in Section 2 of [Additional file](https://zenodo.org/records/12805751) 19). Indeed, the AD threshold points with highest concordance (assumed as CS ≥ 2.85) between every two pipelines (“corresponding points”) were observed across all levels of resolution and followed a linear trend (r2 ≥ 0.99) in all comparisons (Figure 9C, [Additional file](https://zenodo.org/records/12805751) 19, 22 and 23). Despite the good inter-pipeline concordance (even at low threshold levels), differences in the discriminatory power were observed, as shown by deviations from a *y=x* scenario (Figure 9D). In particular, most differences were seen when SeqSphere was involved, as it revealed a higher resolution across all threshold levels. A fine-tuned analysis about pipeline performance and comparability at “outbreak” level is presented below (Section 4.4).

**Figure 9.**
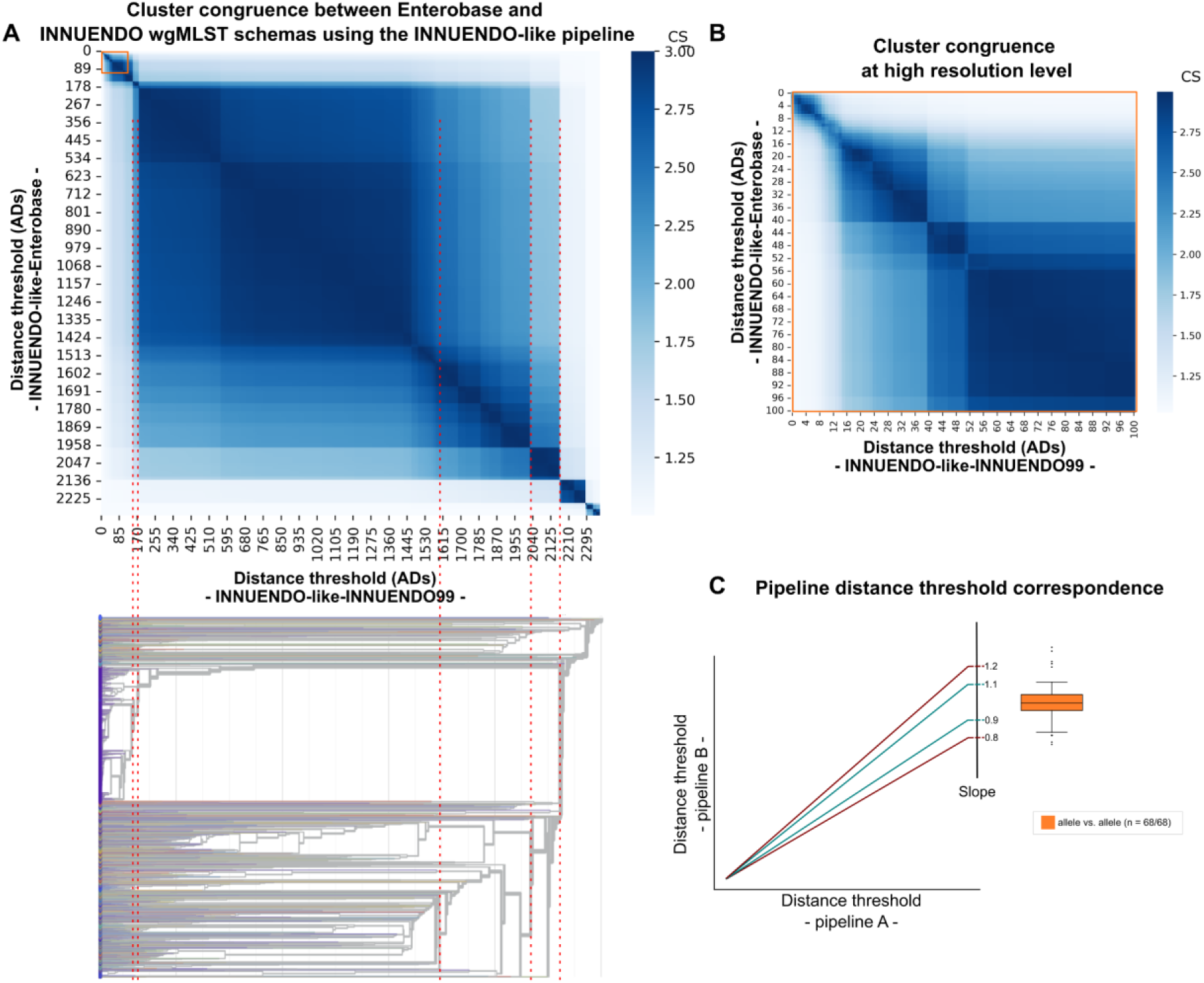
Global evaluation of cluster congruence between different pipelines at all threshold levels for *E. coli* dataset. **A)** Heatmap with the CS obtained for the comparison of the clustering results obtained at all possible thresholds of two pipelines (details on each pairwise comparison are in [Additional file](https://zenodo.org/records/12805751) 19, with INNUENDO-like-Enterobase vs. INNUENDO-like-INNUENDO99 using the HC algorithm being presented here as an example). The orange square highlights the levels of high resolution. To better link the heatmap with the clustering tree, we present an inverted dendrogram (i.e., from the highest to the lowest resolution) showing how the congruence at all levels is related with the phylogenetic structure of the dataset (dendrogram obtained with INNUENDO-like-INNUENDO99 and visualized in auspice.us ^129^). Dashed red lines connect the CS score and clustering results obtained at similar distance thresholds. **B)** Zoom-in in the high resolution level highlighted in orange in panel A. **C)** Representation of the linear trend lines expected for the correspondence points with a theoretical slope deviation of 10% and 20% to be used as scale reference for the boxplots. The slope is used as a proxy to evaluate whether two pipelines yield highly concordant clustering at the exact same level with slopes close to 1 (i.e. *y=x* scenario) reflecting high clustering concordance and similar discriminatory power across all levels. The boxplot presents the slope distribution for allele vs. allele (orange) pipeline comparisons for the linear trendlines with a r2 ≥ 0.99, illustrated in [Additional file](https://zenodo.org/records/12805751) 19 and detailed in [Additional file](https://zenodo.org/records/12805751) 23 (“n” refers to the number of comparisons with r2 ≥ 0.99 over the total number of comparisons). The boxplot of the SNP vs. SNP scenario is not presented as we only had one comparison, and the one of the allele vs. SNP scenario is not presented as only 11/34 comparisons yielded an r2 ≥ 0.99 ([Additional file](https://zenodo.org/records/12805751) 23).

Regarding the SNP-based pipelines, the analysis was conducted with a focus on the clustering obtained at up to a 100 SNPs threshold for the O157:H7 serotype. In summary, CSI Phylogeny provided a higher resolution than SnippySnake (Section 3 of [Additional file](https://zenodo.org/records/12805751) 19), but this observation should not be extrapolated without taking into consideration that the former excluded almost 20% of the sequences. When comparing allele- and SNP-based pipelines (see heatmaps in Section 4 of [Additional file](https://zenodo.org/records/12805751) 19), we again observed a bias towards the higher resolution of CSI Phylogeny, while SnippySnake provided a similar resolution power as the allele-based pipelines.

#### 4.4 Concordance for outbreak detection

Allele-based approaches are the most commonly applied for *E. coli* outbreak detection, but the method and distance threshold used to determine a possible outbreak-related cluster usually varies between laboratories and the inclusion criteria for outbreak is usually set during investigation. The INNUENDO project proposed a dynamic threshold of 0.34% of the cgMLST schema, corresponding to 8 ADs in the INNUENDO cgMLST schema, due to its good concordance with clusters of epidemiologically verified isolates ^20^. We used this 0.34% threshold (which translates into 9 ADs in all pipelines) to start exploring the pipeline congruence at potential outbreak level.

Each pipeline detected between 169 to 182 clusters at 9 ADs, from which, on average, 96.6% had similar composition in at least two pipelines and 3.2% were exclusively detected by a single pipeline ([Additional file](https://zenodo.org/records/12805751) 24). On average, 70.4% of the clusters detected by a given pipeline were also detected with the exact same composition by all remaining pipelines. We further evaluated the minimum threshold level (ADs or SNPs) at which each 9 AD cluster would be detected by the other pipelines. This analysis yielded a total of 185 clusters that, once detected at 9 ADs threshold by at least one pipeline, were detected by all pipelines regardless of the threshold ([Additional file](https://zenodo.org/records/12805751) 25), with SeqSphere and INNUENDO-like-ENTEROBASE requiring slightly higher thresholds to yield the same clusters as the other pipelines. Regardless of this observation, most of the outbreak clusters were detected at a threshold ≤ 9 ADs in all allele-based pipelines, or at higher threshold levels close to 9 ADs (Figure 10A). At SNP level, the assessment was restricted to those “outbreak” clusters corresponding to O157:H7 (94 out of the 185 clusters) and to the SnippySnake pipeline, because of the CSI Phylogeny behavior noticed above. As anticipated above, SnippySnake revealed a density profile similar to allele-based pipelines (Figure 10A), thus demonstrating that core SNP and cg/wgMLST analyses have equivalent performance for O157:H7 outbreak detection, in line with a previous observation ^74^. The difference between the AD thresholds required by the different allele-based pipelines to detect each “outbreak-level” cluster had a median of 3 ADs, with a minimum of 0 and maximum of 15 ADs. This trend is less influenced by the clustering algorithm rather than the cg/wgMLST schema (Figure 10B). A slightly higher diversity was observed between those pipelines relying on the Enterobase schema (median of 2 ADs) than between the ones relying on INNUENDO (median of 1 AD), possibly due to the fact that the Enterobase schema was run with different allele callers (and also different versions of the same allele caller), while all pipelines relying on the INNUENDO schema used chewBBACA (the allele caller used for its refinement) ^72,75^. A side observation was that the direct use of chewBBACA on the Enterobase schema systematically led to around 2% of loci not called in each sample, even though this did not substantially affect the congruence and outbreak detection performance ([Additional file](https://zenodo.org/records/12805751) 19). Looking at pairwise comparisons between all pipelines, our results showed that the overlap of clusters detected at 9 ADs with the exact same composition was 83.7%, on average, a value that increased to 94.9% when applying a flexible threshold according to the median estimation above (i.e. 3 ADs above, Figures 10C and 10D, [Additional file](https://zenodo.org/records/12805751) 26). This exercise further corroborated the outlier behavior of SeqSphere and INNUENDO-like-ENTEROBASE, which needed higher thresholds to yield the same clusters as the other pipelines (Figure 10C, [Additional file](https://zenodo.org/records/12805751) 25 and [Additional file](https://zenodo.org/records/12805751) 26).

**Figure 10.**
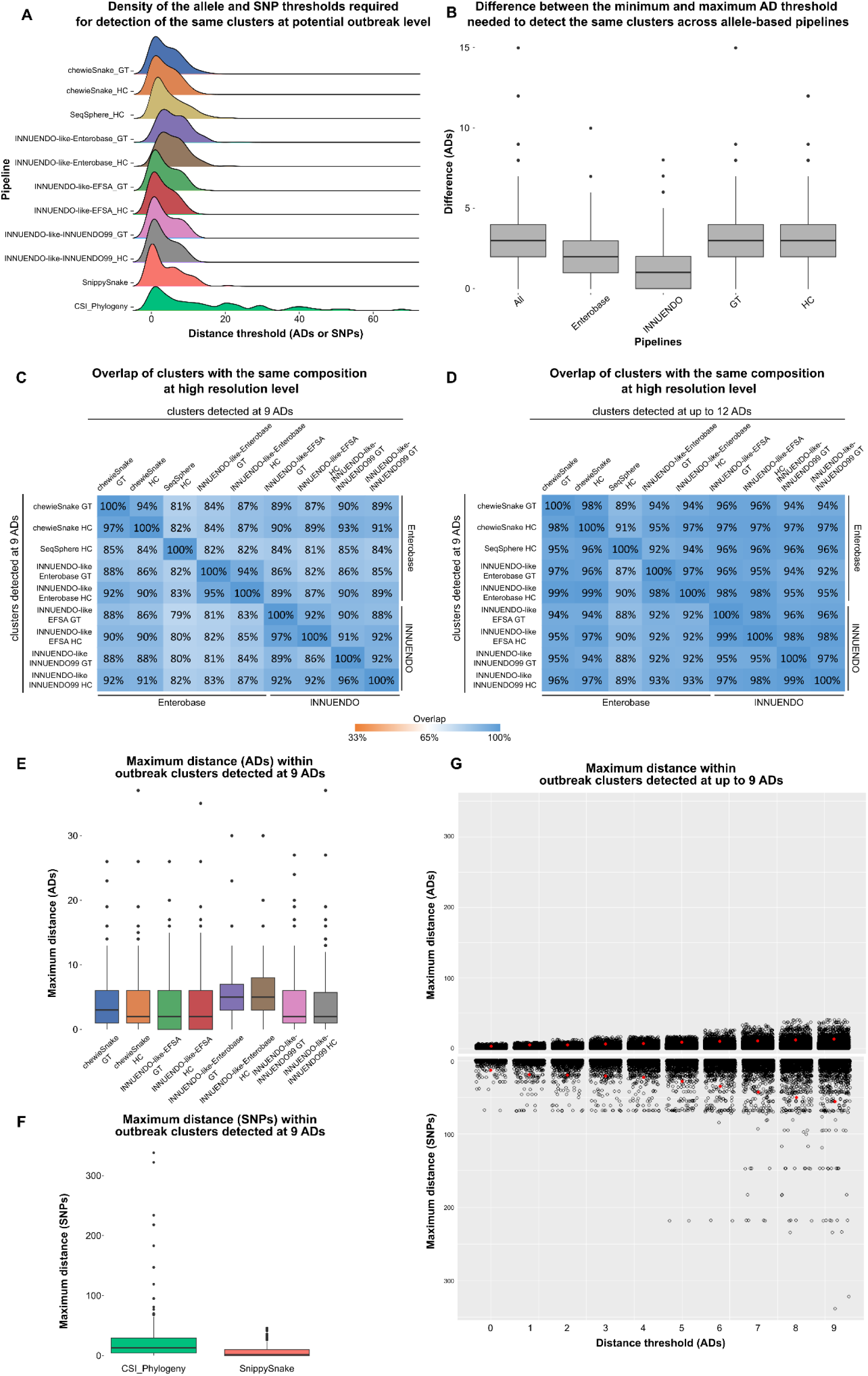
Evaluation of the performance of the genome-scale pipelines in the identification of potential outbreak-related isolates for the *E. coli* dataset. **A)** Density of the distance thresholds (ADs for allele-based pipelines and SNPs for SNP-based pipelines) required for the identification of clusters detected by at least one allele-based pipeline at 9 ADs. Only clusters having the same composition in all allele-based pipelines were included (n = 185). The results of SNP-based pipelines only include the clusters corresponding to the O157:H7 serotype and were obtained using a serotype-specific reference. **B)** Distribution of the difference between the minimum and maximum AD threshold needed to detect the same clusters across allele-based pipelines, using the same clusters as described in panel A. **C)** Overlap between the genetic clusters detected by allele-based pipelines at 9 ADs. **D)** Overlap between the genetic clusters detected by one pipeline at 9 ADs and those detected by the others at ≤ 12 ADs. **E)** Distribution of the maximum ADs within each “outbreak-level” cluster identified at 9 ADs by the respective pipeline. **F)** Distribution of the maximum SNP distances within “outbreak-level” cgMLST clusters identified by at least an allele-based pipeline at 9 ADs. **G)** Maximum allele (top) or SNP (bottom) distance threshold within each cluster determined by at least an allele-based pipeline at up to 9 ADs. The red dot indicates, for each threshold, the 95^th^ percentile.

When looking at the genomic diversity (SNPs/ADs) within the cg/wgMLST “outbreak clusters” (9 ADs), the INNUENDO-like-ENTEROBASE consolidated its outlier behavior by capturing a higher genomic diversity within the clusters and showing higher intra-cluster maximum distances with incremental cluster sizes than the other pipelines (Figures 10E and 10F). The other outlier pipeline, SeqSphere, could not be used for this particular analysis, due to the unavailability of suitable pairwise distance input within the BeONE project. SnippySnake again provided similar clustering and outbreak resolution power as the allele-based pipelines (Figure 10E). For example, 95% of the clusters detected at 5 ADs by at least one allele-based pipeline were composed by strains that diverge by no more than 9 alleles, a value that only slightly increased to 12 when assessed in terms of SNPs (Figure 10G).

Finally, we conducted an additional exercise with the pipeline running an wgMLST schema (INNUENDO-like-INNUENDO99) to explore the potential gain in resolution to discriminate potential outbreak isolates (as assessed by cgMLST) when increasing the number of wgMLST loci under comparison, aligned with a previously explored rationale ^20,55^. Regardless of the clustering algorithm, this approach resulted in an average increase of 5 ADs in the maximum pairwise distances observed between the isolates of the same original cgMLST cluster ([Additional file](https://zenodo.org/records/12805751) 27), demonstrating the clear increase in resolution provided by the dynamic extension of the cgMLST schema with wgMLST loci shared by the same-outbreak isolates.

### 5. Campylobacter jejuni

*Campylobacter jejuni* dataset (Figure 11A) was analyzed with seven allele-based and one SNP-based pipeline (Table 1 and Tables S1.1 and S1.2 in Additional file 28).

**Figure 11.**
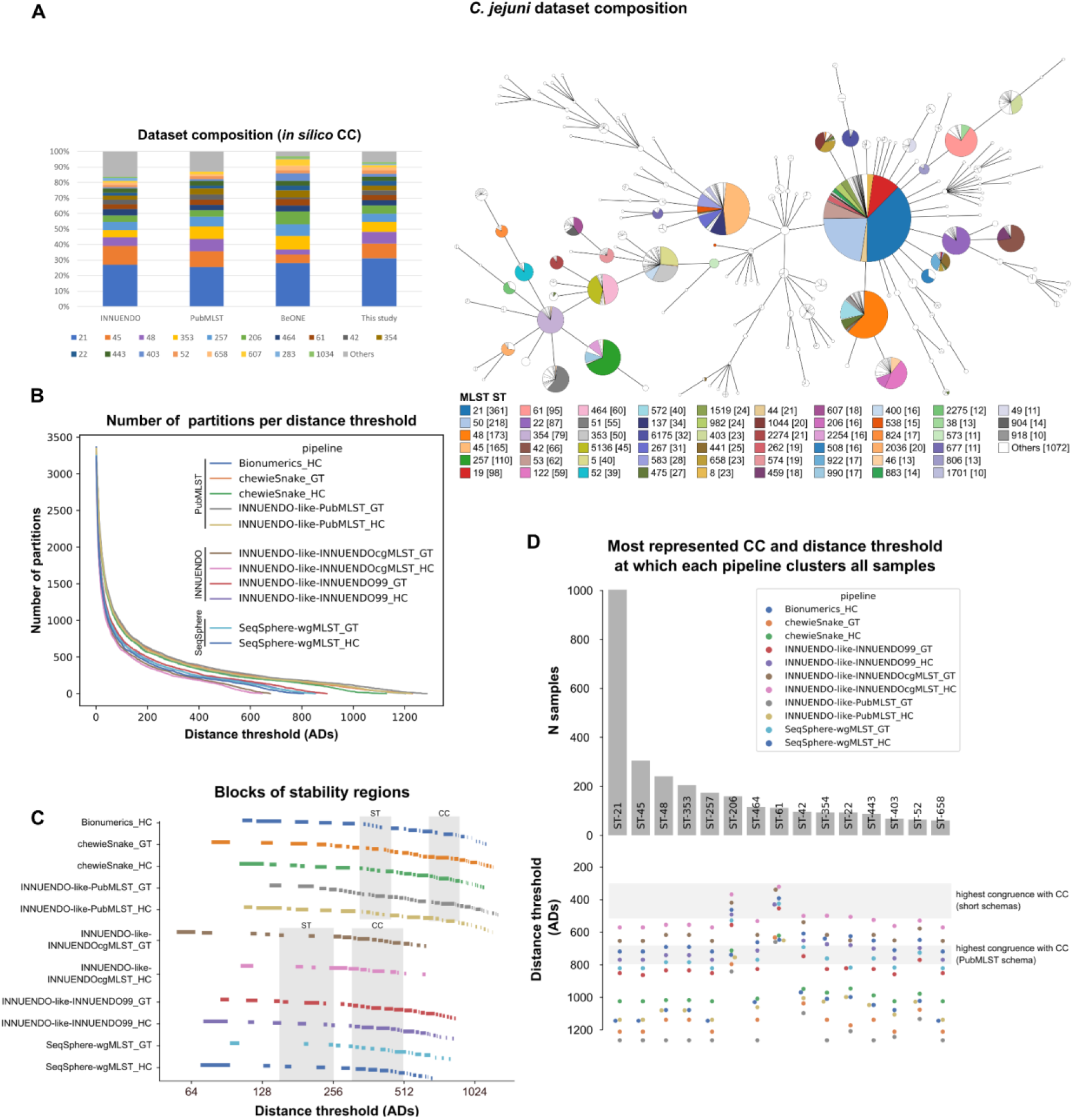
Assessment of allele-based clustering at all possible threshold levels for *C. jejuni* and congruence with traditional serotype. **A)** Composition of the *C. jejuni* dataset used in this study in terms of CC and in comparison with the composition of the datasets INNUENDO ^20^ and the PubMLST database as of November 2021 ^21^. A GrapeTree ^56^ visualization of the MST obtained with the INNUENDO-like-PubMLST pipeline is shown. Nodes (i.e. samples) are collapsed at the threshold with highest congruence with CC (839 ADs for this pipeline) and colored according to the ST classification. **B)** Number of partitions obtained by each pipeline at each possible distance threshold. **C)** Clustering stability regions determined for each pipeline. The different blocks of each pipeline start in a different line. Distance thresholds (*x* axis) are presented in log2 scale. **D)** Barplot (top) with the number of samples of the top represented CCs (≥ 50 samples) in *C. jejuni* dataset, with a swarmplot (bottom) indicating the AD threshold at which each pipeline clusters together all samples of each CC.

**Figure 12.**
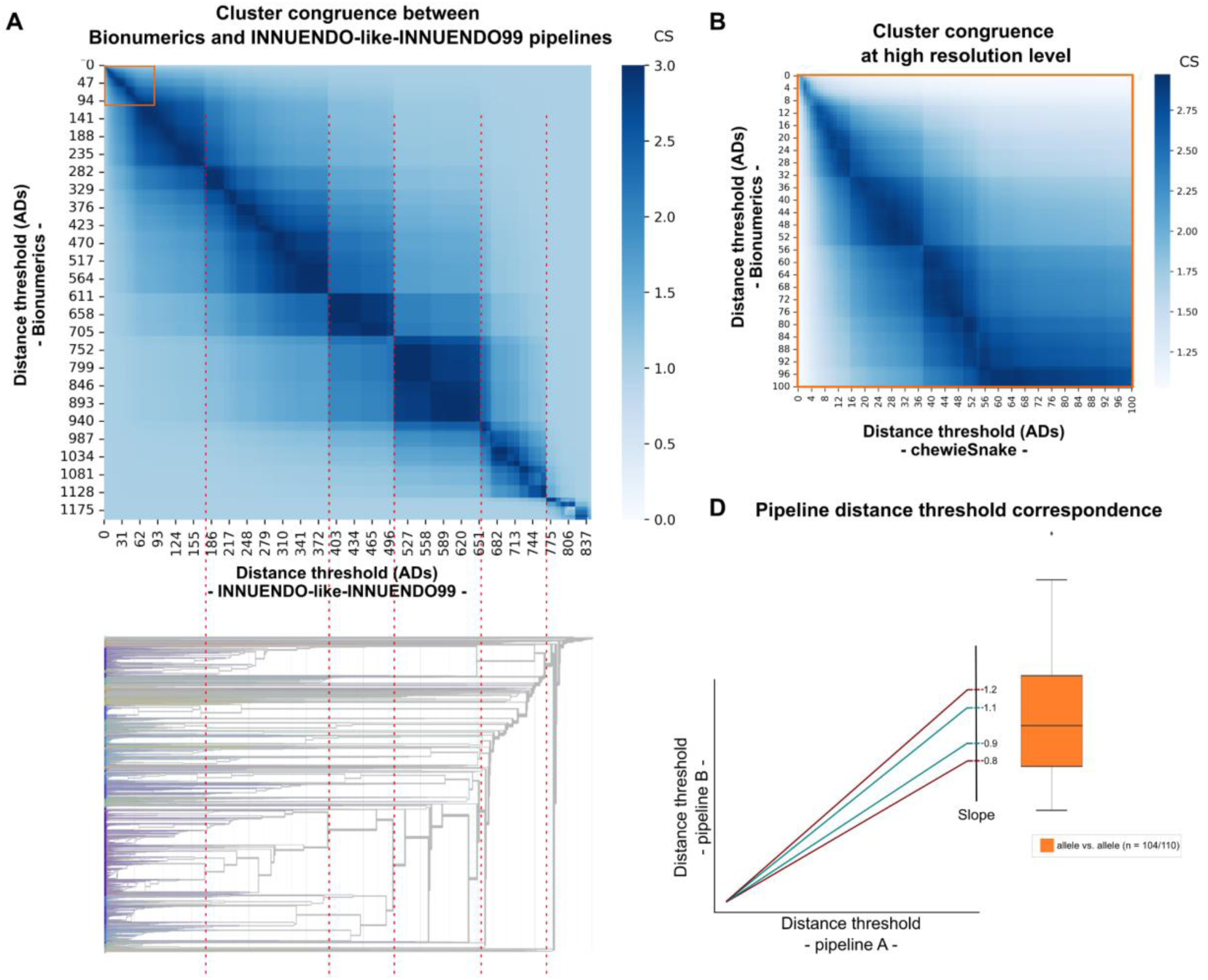
Global evaluation of cluster congruence between different pipelines at all threshold levels for *C. jejuni* dataset. **A)** Heatmap with the Congruence score (CS) obtained for the comparison of the clustering results obtained at all possible thresholds of two pipelines (details on each pairwise comparison are in [Additional file](https://zenodo.org/records/12805751) 28, with Bionumerics vs. INNUENDO-like-INNUENDO99 using the single-linkage hierarchical clustering (HC) algorithm being presented here as an example). The orange square highlights the levels of high resolution. To better link the heatmap with the clustering tree, we present an inverted dendrogram (i.e., from the highest to the lowest resolution) showing how the congruence at all levels is related with the phylogenetic structure of the dataset (dendrogram obtained with INNUENDO-like-INNUENDO99 and visualized in auspice.us ^129^). Dashed red lines connect the CS score and clustering results obtained at similar distance thresholds. **B)** Zoom-in in the high resolution level highlighted in orange in panel A. **C)** Representation of the linear trend lines expected for the correspondence points with a theoretical slope deviation of 10% and 20% to be used as scale reference for the boxplots. The slope is used as a proxy to evaluate whether two pipelines yield highly concordant clustering at the exact same level with slopes close to 1 (i.e. *y=x* scenario) reflecting high clustering concordance and similar discriminatory power across all levels. The boxplot presents the slope distribution for allele vs. allele (orange) pipeline comparisons for the linear trendlines with a r2 ≥ 0.99, illustrated in [Additional file](https://zenodo.org/records/12805751) 28 and detailed in [Additional file](https://zenodo.org/records/12805751) 32 (“n” refers to the number of comparisons with r2 ≥ 0.99 over the total number of comparisons). The boxplots of the SNP vs. SNP and allele vs. SNP scenarios are not presented as only 4/14 and 6/160 comparisons yielded an r2 ≥ 0.99 ([Additional file](https://zenodo.org/records/12805751) 32).

#### 5.1 Evaluation of allele-based clustering and comparison of stability regions

Following QC of the initial 3686 *C. jejuni* isolates ([Additional file](https://zenodo.org/records/12805751) 1), the pipelines using the large and static cgMLST schema from PubMLST (1343 loci) ^76^ retained ∼95% of the samples, while the ones with shorter cgMLST schemas retained ∼98.5% (Table S1.1 in [Additional file](https://zenodo.org/records/12805751) 28). The later either perform cgMLST on a set of core loci dynamically extracted from an wgMLST schema (either 899/2754 core loci of the INNUENDO schema ^20^ or 860/1595 core loci of the Ridom schema ^77^) for this dataset, or run a short and static cgMLST (678/2754 core loci of the INNUENDO schema ^20^) (Table S1.1 in [Additional file](https://zenodo.org/records/12805751) 28). These three pipeline groups, relying on different schema sizes, revealed distinct clustering patterns in terms of number of clusters across all possible thresholds (Figure 11B), with the pipelines using PubMLST schema displaying a higher resolution, i.e. a higher number of clusters for the same threshold.

Independently of the schema and clustering algorithm (GT or HC), no pipeline revealed regions of high stability until 55 ADs, far above the common thresholds for outbreak detection. After this point, although multiple regions of high stability have been identified across all levels, they were quite small, hampering the identification of stability plateaus common to all pipelines (Figure 11C). This scenario contrasts with the other species analyzed in this study and likely reflects the high genetic diversity, extensive mosaicism and polyclonal population structure of *C. jejuni* ^78–80^.

#### 5.2 Evaluation of allele-based clustering congruence with traditional typing and WGS-derived pathogen main lineages

Regarding the cg/wgMLST clustering congruence with CC classification, the pipelines relying on PubMLST schema presented the highest congruence point between 644 and 839 ADs (CS∼2.7), while this threshold ranged between 315 and 522 (CS ∼2.7) for the pipelines with shorter schemas (Table S1.1 in [Additional file](https://zenodo.org/records/12805751) 28). Still, the clustering of each pipeline at this point yielded a similar number of partitions, which varied between 161 and 229. When assessing the distribution of the 39 CCs across these partitions, only 25% had all the respective samples integrated in the same cluster. In another perspective, almost two thirds of the CCs have strains dispersed across three or more clusters at this level. Despite the fact that the likelihood of two samples of the same CC cluster together at this cgMLST level is still high (AWC > 0.8 for all pipelines), our results show that the CC classification only slightly mimics *C. jejuni* clustering at genome scale. Indeed, the thresholds needed to cluster all samples of the same CC are quite high across all pipelines, almost reaching the size of the respective cgMLST schema, as observed for most of the dominant CCs in this dataset (Figure 11D and [Additional file](https://zenodo.org/records/12805751) 29). The comparison between cgMLST clustering and ST classification revealed a more informative scenario, as 78.2% of the 227 STs present in this dataset have all their samples grouped into a single cluster at the highest congruence point with cgMLST (Table S1.1 in [Additional file](https://zenodo.org/records/12805751) 28), and only 4 to 7% of the STs were split into three or more clusters at this level. Moreover, it allowed the identification of STs with very different genetic heterogeneity in the dataset. For instance, when examining the most prevalent STs in the dataset (Figure 11D), some (e.g. ST45, ST48 and ST50) exhibited significant intra-ST diversity, which escalates with the cgMLST schema size (as seen in the CC evaluation). In contrast, others displayed less diversity, requiring similar threshold levels for merging all samples from the same ST, regardless of the pipeline used ([Additional file](https://zenodo.org/records/12805751) 30 and Figure S1.1 in [Additional file](https://zenodo.org/records/12805751) 28).

Finally, we assessed the congruence between cg/wgMLST clustering and WGS-derived pathogen main lineages inferred through PopPUNK ^73^. For this *C. jejuni* dataset, PopPUNK clustering had the highest congruence with cg/wgMLST typing (CS ∼2.6) at allelic distance thresholds consistently falling in between the points of highest congruence with ST and CC (Table S1.1 in [Additional file](https://zenodo.org/records/12805751) 28).

#### 5.3 Evaluation of cluster congruence between different pipelines at all threshold levels

Our in-depth pairwise congruence analysis showed a high concordance between all allele-based pipelines (as exemplified for a pairwise comparison in Figures 11A and 11B, and detailed in Section 2 of [Additional file](https://zenodo.org/records/12805751) 28). Indeed, the AD threshold points with highest concordance (assumed as CS ≥ 2.85) between every two pipelines (“corresponding points”) followed a linear trend (r2 ≥ 0.988) in all comparisons (Figure 11C, [Additional file](https://zenodo.org/records/12805751) 28, 31 and 32). Not unexpectedly, due to the differences in cgMLST schema size between pipelines, the discriminatory power was consistently higher in the PubMLST schema pipelines, as shown by deviations from a *y=x* scenario (Figure 6D). Moreover, the pairwise comparisons between these pipelines revealed many corresponding points even within the “outbreak region” ([Additional file](https://zenodo.org/records/12805751) 31), which was not observed in the comparisons involving the pipelines with shorter schemas. A fine-tuned analysis about pipeline performance and comparability at “outbreak” level is presented below (Section 5.4). The pairwise comparisons between allele-based pipelines and the available SNP-based pipeline (CSI Phylogeny) was conducted with a focus on the clustering obtained at up to a 100 SNPs threshold for the top represented STs, namely ST21, ST50, ST45, ST48 and ST257. Our results showed that CSI Phylogeny with an ST-specific reference generally offered superior resolution compared to allele-based pipelines using short schemas, although less frequently than when comparing with those employing PubMLST schemas ([Additional file](https://zenodo.org/records/12805751) 32).

#### 5.4 Concordance for outbreak detection

Allele-based approaches are expected to become the standard method for *C. jejuni* outbreak detection, but this is not yet conducted routinely in most countries. Despite the existence of limited data about the cgMLST resolution levels with good epidemiological concordance, a threshold of 4 ADs has been often pointed out as a proxy for generating outbreaks signals ^81^, with proven application in real investigations ^81,82^. As such, in the present study, in order to start exploring the pipeline congruence at potential outbreak level, we applied a 4 AD threshold for all pipelines.

Each pipeline detected between 271 to 388 clusters at 4 ADs, from which, on average, 97.1% had similar composition in at least two pipelines and 2.9% were exclusively detected by a single pipeline ([Additional file](https://zenodo.org/records/12805751) 33). However, as anticipated above, due to the substantially different sizes of the studied cgMLST schemas, the clustering congruence at a 4 AD threshold was considerably higher between pipelines with similar schema sizes (Figures 13A and 13B). On average, only 36.6% of the clusters detected by a given pipeline were detected with the exact same composition in all pipelines, but this value considerably increased when this comparison is restricted to pipelines with similar cgMLST schema size (81.7% for pipelines running the PubMLST schema and 58.6% for the others) ([Additional file](https://zenodo.org/records/12805751) 33). We subsequently sought to evaluate the minimum threshold level (ADs or SNPs) at which each 4 AD cluster would be detected by the other pipelines. This analysis yielded a total of 430 clusters that, once detected at 4 ADs threshold by at least one pipeline, were detected by all pipelines regardless of the threshold ([Additional file](https://zenodo.org/records/12805751) 34). The cg/wgMLST pipelines with larger schema very often needed thresholds two or three times higher to provide the same clusters, which reflects their higher resolution power ([Additional file](https://zenodo.org/records/12805751) 34). When looking at this threshold dispersion in terms of SNPs (with the CSI Phylogeny), most of the cgMLST clusters at 4 ADs were detected at a threshold ≤ 4 SNPs or at higher threshold levels close to 4 SNPs (Figure 13A). The difference between the AD thresholds required by the different allele-based pipelines to detect each “outbreak-level” cluster had a median of 3 ADs, with a minimum of 0 and maximum of 24 ADs. This trend is less influenced by the clustering algorithm than the cg/wgMLST schema, as a median of only 1 AD difference is observed when comparing pipelines running schemas with similar sizes (Figure 13B). Looking at pairwise comparisons between all pipelines, our results showed that the overlap of clusters detected at 4 ADs with the exact same composition was, on average, 88.6% between pipelines using the PubMLST schema and 75.0% between the pipelines running the shorter schemas (Figure 13C and [Additional file](https://zenodo.org/records/12805751) 35). Importantly, given the significant difference in resolution at outbreak level, one would need to decrease the threshold applied in the shorter schema pipelines to a threshold as low as 1 AD to have a proxy of the potential outbreak clusters obtained with the PubMLST schema at the commonly used 4 ADs threshold (Figure 13D and [Additional file](https://zenodo.org/records/12805751) 35). This hampers the application of a single static threshold across different pipelines, highlighting the challenge of applying shorter schemas for outbreak detection. Given these results, we exclusively measured the genomic diversity within potential outbreak level clusters (i.e, 4 ADs) for the pipelines using the PubMLST cgMLST schema. The maximum distance between same-cluster samples per pipeline had an average of 2 ADs, a value that increases to 3 SNPs when mapping to an ST-representative reference (Figure 13E).

**Figure 13.**
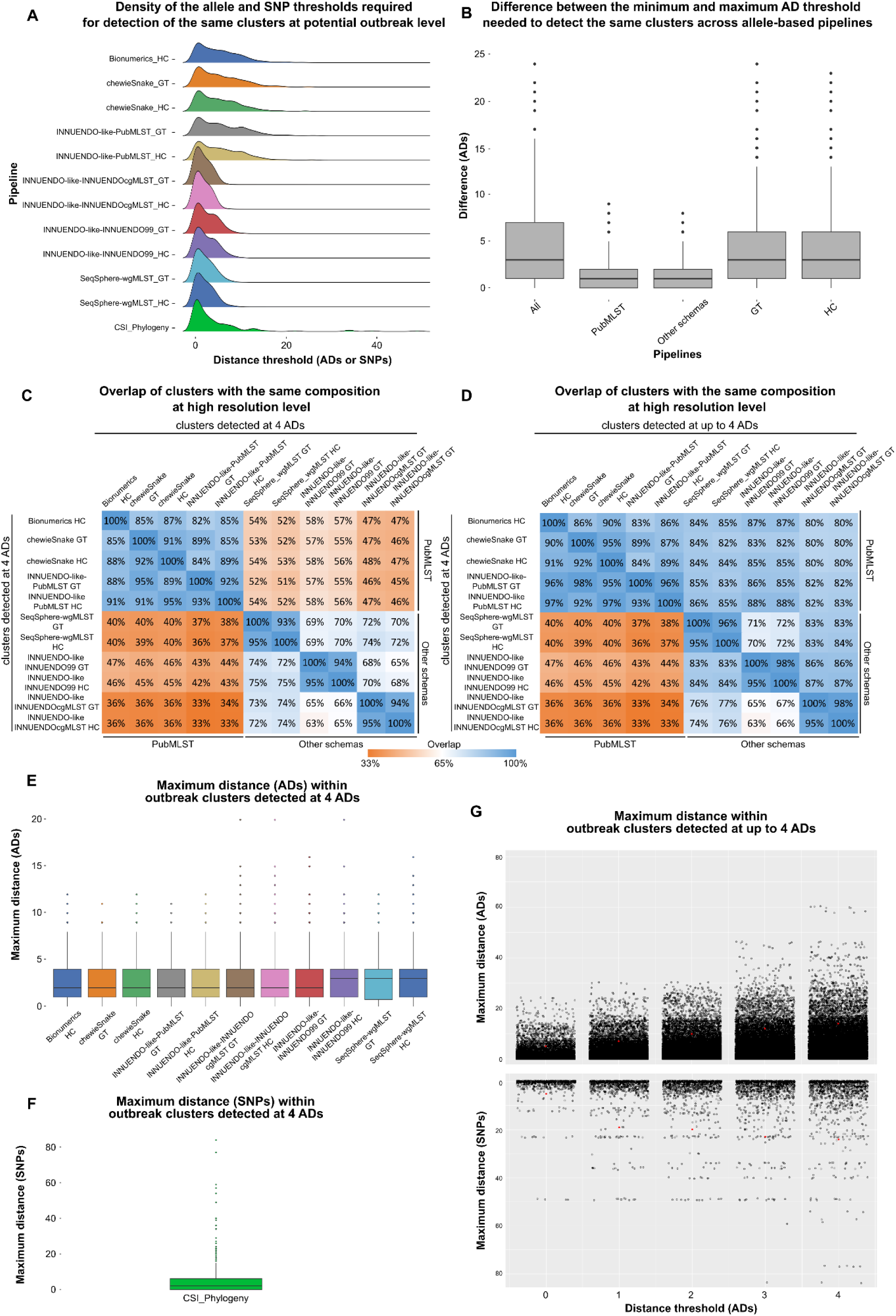
Evaluation of the performance of the genome-scale pipelines in the identification of potential outbreak-related isolates for the *C. jejuni* dataset. **A)** Density of the distance thresholds (ADs for allele-based pipelines and SNPs for SNP-based pipelines) required for the identification of clusters detected by at least one allele-based pipeline at 4 ADs. Only clusters having the same composition in all allele-based pipelines were included (n = 430). The results of the SNP-based pipeline only include the clusters corresponding to the top 5 STs and were obtained using an ST-specific reference. **B)** Distribution of the difference between the minimum and maximum AD threshold needed to detect the same clusters across allele-based pipelines, using the same clusters as described in panel A. **C)** Overlap between the genetic clusters detected by allele-based pipelines at 4 ADs. **D)** Overlap between the genetic clusters detected by one pipeline at 4 ADs and those detected by the others at ≤ 4 ADs. **E)** Distribution of the maximum ADs within each “outbreak-level” cluster identified at 4 ADs by the respective pipeline. **F)** Distribution of the maximum SNP distances within “outbreak-level” cgMLST clusters identified by at least an allele-based pipeline at 4 ADs. **G)** Maximum allele (top) or SNP (bottom) distance threshold within each cluster determined by at least an allele-based pipeline at up to 4 ADs. The red dot indicates, for each threshold, the 95^th^ percentile.

Finally, we conducted an additional exercise to evaluate the resolution gain obtained when applying a previously explored rationale ^20,55^ in which the cgMLST is dynamically increased to the maximum number of wgMLST shared loci for a given cgMLST cluster. This exercise was conducted for the wgMLST INNUENDO schema (N = 2754 loci), starting either from a static core of 678 loci (INNUENDO-like-INNUENDOcgMLST) or from a core of 899 loci shared by 99% of the studied dataset (INNUENDO-like-INNUENDO99), as well as for the SeqSphere wgMLST schema (N = 1595 loci), starting from a core of 860 loci shared by 99% of the studied dataset (SeqSphere-wgMLST). In all cases, there was a clear gain in the discriminatory power, with an increase of about 5 ADs in the maximum pairwise distances observed between the isolates of the same original cgMLST cluster ([Additional file](https://zenodo.org/records/12805751) 36). These clusters often enroll isolates with considerably high genetic distances ([Additional file](https://zenodo.org/records/12805751) 36), thus consolidating the observation that a threshold of 4 ADs is high for pipelines with original short cgMLST schemas, as it would likely overestimate the size of a real outbreak. The application of a dynamic wgMLST approach as the one tested here might mitigate this risk, providing a layer of extra resolution towards a more reliable detection of outbreak signals.

### 6. Impact of applying a different assembly pipeline and updating the allele-caller: proof-of-concept study

Given the availability of an alternative assembly pipeline within the Consortium (INNUca ^20,83^), we sought to investigate if its usage in replacement of AQUAMIS would impact the study outcomes. Additionally, taking into account that the main allele-caller tested in this study (chewBBACA ^72^) was significantly updated while we were conducting our analyses, we extended this proof-of-concept investigation by also comparing the clustering results obtained when two different versions of this software are used, namely the stable chewBBACA v2.8.5 and the recent (at the time of this analysis) chewBBACA v3.3.5. In summary, for each species, we took the INNUENDO-like pipeline (exactly as applied throughout the whole study, i.e. providing the AQUAMIS assemblies as input and using chewBBACA v2.8.5 - details in the Methods section) and compared its clustering results with those obtained when INNUca assemblies or chewBBACA v3.3.5 are used. Similar to the species-specific sections, these exercises were conducted for the two clustering methods used in this study (i.e. single-linkage and MSTreeV2). Our results revealed a high cluster congruence at all threshold levels for all species, with the usage of either INNUca or chewBBACA v3.3.5 yielding barely no deviation from the theoretical *y=x* scenario (slope varying between 0.984 and 1.003), indicating that the clustering at one level with the original pipeline is highly concordant with the clustering at the exact same level in each of the two alternative workflows ([Additional file](https://zenodo.org/records/12805751) 37). We further performed an in depth comparison at outbreak level. Although our pairwise comparison strategy is quite strict, i.e. only considers a cluster as detected by two pipelines if it has exactly the same composition at the same AD level, the alternative pipelines were able to detect more than 94% and 97% of the clusters yielded by the original pipeline, when INNUca assemblies or chewBBACA v3.3.5 are used, respectively ([Additional file](https://zenodo.org/records/12805751) 37). These values are considerably higher than those obtained in the inter-pipeline comparisons presented above for each species. Altogether, this exercise shows that the species-specific results and conclusions of this study would be maintained if other assemblies have been provided or if the partners using chewBBACA had updated their respective pipeline to the newest version of this software. In another perspective, it underscored the added value of the tools developed in this study to comprehensively and rapidly evaluate the impact of software updates and changes of components, which is pivotal to ensure the long-term robustness and sustainability of routine genomics workflows.

## Discussion

The integration of WGS into foodborne disease surveillance represents a significant advance in public health, providing unparalleled resolution to detect and respond to outbreaks. Still, the gains of WGS can only be fully leveraged by taking a One Health approach, which is not free of challenges ^5^. For instance, intra-country and supra-national One Health surveillance enrolls multiple stakeholders (commonly in different stages of WGS application) and different surveillance systems (decentralized and centralized), complicating harmonization and communication ^5,40,84^. Moreover, with the multitude of bioinformatics pipelines available nowadays, which are under constant innovation and refinement, it is now clear that, even though cg/wgMLST are becoming the standard WGS typing methods for FWD outbreak detection and investigation, the community is not evolving towards the adoption of a common WGS pipeline, as demonstrated in our study. In this context, as reinforced by WHO, it is critical to guarantee that WGS solutions are comparable between laboratories at regional, national and international levels ^13^. In the present study, we relied on a collaborative effort of multiple European institutes from seven different Countries, from the food, animal and human health sectors, to perform a large-scale assessment of the comparability and congruence at clustering level of different WGS pipelines for four major foodborne bacterial pathogens: *L. monocytogenes*, *S. enterica*, *E. coli*, and *C. jejuni*. We took a surveillance-guided approach in which each participating institute ran their pipeline over the same sequencing dataset, rather than conducting a theoretical comparison focused on assessing the impact of individual software components on the overall pipeline performance. This strategy provided a more realistic picture of how the clustering results obtained by different laboratories in their routine activities can be compared between each other at all possible resolution levels, while unveiling the WGS typing congruence with traditional methods and the inter-pipeline performance for outbreak detection.

In this comprehensive study, we observed variations in clustering patterns across different bioinformatics pipelines, reflecting the diversity of approaches being currently applied for routine genomic analysis. Still, we generally found an overall good concordance for the pipelines following a cg/wgMLST approach, which displayed a high clustering congruence and similar discriminatory power across all levels of resolution, with the notable exception of *C. jejuni* comparisons. Indeed, for this pathogen, we observed a significant variation in resolution power that could be linked to the inclusion of pipelines running over schemas with a very discrepant number of loci. While this discrepancy reflects the current lack of a standard schema for *C. jejuni*, our observation that cgMLST pipelines with larger schemas often required thresholds two or three times higher than those with shorter schemas to detect the same clusters raises concerns about the lower performance (or even reliability) of the latter in discriminating outbreak clusters. In this regard, our results suggest that the application of a dynamic wgMLST approach that takes advantage of the accessory genome ^57–59^ might mitigate this risk, as simulated here for *C. jejuni, S. enterica* and *E. coli*.

Another significant observation was that even allele-based pipelines with overall high congruence exhibited some non-negligible differences in detecting outbreak clusters. Although our comparative analysis at outbreak level was intentionally strict, only considering clusters with the same isolate composition as concordant between pipelines, it was crucial to uncover discrepancies with potential practical impact on outbreak case definition and inter-laboratory communication. Indeed, despite the case-definition criteria typically include a static and similar threshold for both centralized (e.g. ECDC and EFSA) and national pipelines ^36–39^, our findings indicate that a cut-off flexibilization by up to 2-3 ADs increases the likelihood that a given pipeline detects clusters with the exact same composition as another pipeline by roughly 10%. Consistently with previous observations ^41^, our results also showed that same-schema pipelines tend to detect the same outbreak signals at more similar thresholds, and that, in this scenario, the flexibilization of just 1 AD is often sufficient to reach very concordant performance in outbreak detection. Importantly, the application of GT or HC clustering methods, which are the most widely applied for cg/wgMLST analysis ^27^, had considerably less impact on pipeline discrepancies. We anticipate that, contrary to the choice of the schema, the current lack of consensus about the clustering method to employ should not be a main factor hampering inter-laboratory comparability. For *L. monocytogenes* and *S. enterica*, we also simulated the application of a more stringent cut-off (e.g., 4 ADs for *L. monocytogenes* and 5 ADs for *S. enterica*), which consolidated that the cut-off flexibilization favors the detection of the same outbreak signal in different laboratories. In practice, as the discrepancy increases with the variability of pipelines under comparison, a potential reasonable approach when setting international and inter-sectoral case definition criteria can be the inclusion of suspected outbreak samples based on a flexible threshold, instead of the usual reliance on a static and similar cut-off that does not take into account the comparability of the pipelines involved in the investigation. In the context of a multi-national outbreak, the proposed approach of relaxing and tailoring the outbreak cut-off would reduce the likelihood of missing isolates that go unreported as they fall outside the case definition threshold in a given national pipeline, but that would cluster within the defined threshold in the pipeline centralizing the genomic outbreak investigation (e.g. ECDC or EFSA).

The adoption of this strategy for outbreak case-definition implies shaping the threshold boundaries according to the specific context of the outbreak under investigation, namely: i) the genetic and temporal distance between the outbreak isolates already with a confirmed epidemiological link; ii) the clustering performance and comparability of the pipelines involved in the outbreak investigation (regional, national or supra-national); and, iii) the expected genetic heterogeneity, evolutionary context and available typing information (ST, CC, serotype, drug susceptibility profile) of the potential outbreak-causing strain. While the first premise is expected to be strengthened as the integration of genomic and epidemiological data becomes more frequent (at national and international levels), we consider that the present study adds significant value to better inform outbreak investigation regarding the last two knowledge gaps. On the one hand, we present actual data regarding the comparability of a panoply of pipelines currently in place in several countries and sectors, and describe an innovative methodology for the rapid and comprehensive assessment of pipeline clustering congruence and outbreak performance comparability. On the other hand, with the assessment of the congruence between WGS typing and traditional typing data, we showcase very different levels of genetic heterogeneity of the most prevalent STs, CCs and/or serotypes, which represents valuable information for routine genomic surveillance and outbreak case definition. Indeed, while we consolidated the existence of STs/CCs/serotypes with a clear polyphyletic signature, such as the ST7 and the ST325 for *L. monocytogenes* and the Thompson and the Newport serotypes for *S. enterica,* we also found contrasting scenarios where the intra-ST or intra-serotype diversity was quite low, such as the ST8 and the ST87 for *L. monocytegenes,* the Agona serotype for *S. enterica* and the ST53 for *C. jejuni*. Besides the potential implications that these different evolutionary patterns may have for the selection of isolates for routine WGS surveillance based on traditional typing data, which is commonly performed for the highly prevalent *S. enterica* and *C. jejuni* species ^85^, these observations underscore the potential inadequacy of applying a single “outbreak threshold” within a species, as this may lead to an oversizing of potential outbreak clusters when highly clonal WGS-derived lineages (often overlapping with STs/serotypes) are involved. For instance, a previous in-depth lineage-specific WGS analysis of *L. monocytogenes* evolution also corroborates that the outbreak cut-offs should not disregard the sub-lineages/CCs under analysis ^86^. This topic warrants further research in order to determine which threshold ranges render the highest epidemiological concordance per genogroup or even to evaluate the need of (novel) cg/wgMLST typing schemas tailored to their population structure and evolutionary history. The integration of epidemiological data in large-scale WGS studies, for example, through the analysis of epidemiologically confirmed outbreaks, will be also crucial to tackle this subject.

Noteworthy, even though allele-based pipelines are increasingly consolidated as the method of choice for routine FWD WGS typing with recognized gains ^6–8,20,87^, one cannot dismiss the application of a SNP-based analysis for FWD outbreak detection and investigation. Indeed, they are intuitively used by many laboratories for an enhanced discrimination of outbreak isolates detected by cg/wgMLST, and are still in place in others as main approach for FWD outbreak detection (e.g. SnapperDB or CFSAN SNP pipeline ^16,30,88–90^). The timeframe of this international Consortium and the dimension of our study rendered it impractical to perform an in-depth SNP analysis for each one of the ∼100 to ∼300 potential outbreak clusters consistently detected by all allele-based pipelines for each species. Still, by conducting a SNP analysis for the top represented STs/serotypes, we investigated the congruence between SNP and cg/wgMLST clustering and measured the SNP diversity within potential cg/wgMLST outbreak clusters. Our results suggest that, when using an ST- or serotype-specific reference (similar to what is done in SnapperDB), SNP-based analyses frequently provide equal to higher discriminatory power than cg/wgMLST pipelines, which is also reflected in the higher SNP than allelic distance within a cg/wgMLST outbreak cluster. However, not unexpectedly, this observation was sensitive to the genetic heterogeneity of the dataset, and, consequently, influenced by the ST/serotype under evaluation by the SNP-based pipeline and by the cg/wgMLST schema used by each allele-based pipeline. For example, in *C. jejuni*, although dependent on the ST, ST-specific SNP-based clustering generally provided higher resolution than the allele-based pipelines running shorter schemas, but not so frequently when compared to the pipelines relying on the biggest schema (PubMLST). When comparing SNP-based pipelines between themselves (only possible for *L. monocytogenes* and *S. enterica*), there were noticeable resolution discrepancies, particularly in *S. enterica* analyses. These discrepancies were related with the QC inclusion criteria applied by each SNP-based pipeline, which had a considerable impact on the size of the core SNP alignment, and, consequently, on the resolution. It is noteworthy that our study did not explore the need and benefits of conducting more advanced phylogenetic reconstructions (e.g., Maximum Likelihood) and/or integrating recently described models that incorporate epidemiological and evolutionary data (e.g., substitution rates) ^91^ in SNP-based outbreak detection and investigation. In summary, the variations detected between SNP-based pipelines and their sensitivity to multiple factors showcase the known difficulties in the comparison and integration of surveillance data from laboratories relying on this approach, as often reported in FWDs EQAs ^32–34^. In another perspective, our study suggests that SNP-based pipelines provide enough resolution for outbreak detection when performed at ST level, which consolidates their great potential to leverage even higher resolution when outbreak-specific references are used to zoom-in cg/wgMLST-derived outbreak clusters. Regarding this topic, we explored a dynamic cg/wgMLST approach that performs such zoom-in by automatically increasing the resolution through the inclusion of shared accessory wgMLST loci ^20,55^, as implemented in ReporTree ^55^. Although we did not perform a direct comparison to assess the congruence between the two zoom-in approaches, the results of this exercise sustain that the “cg/wgMLST only” approach can be a promising alternative to enhance the intra-outbreak resolution, while avoiding the need of relying on different methodologies and likely reducing the complexity and turn-around time of the genetic investigation in the context of outbreak.

The relevance of large-scale WGS typing approaches goes far beyond the detection and investigation of outbreaks. Indeed, the identification and real-time monitoring of main circulating lineages is pivotal for a sustainable and efficient pathogen surveillance. Therefore, in the last few years, a significant effort has been made towards the development of clustering-based nomenclatures. For instance, for cgMLST, Enterobase and INNUENDO propose nomenclature systems based on the hierarchical clustering at different resolution levels ^20,92^, and other approaches relying on Life Identification Numbers (LIN) were recently released ^93,94^. For SNPs, the hierarchical “SNP-address” nomenclature has proven applicability in routine surveillance. In this study, we could identify, for each species and pipeline, subsequent distance thresholds in which cluster composition remains similar (i.e. “stability regions”). Despite slight deviations, concordant regions were normally found across all pipelines, thus showcasing their suitability to capture main WGS-derived lineages and species population structure. For instance, *L. monocytogenes, S. enterica* and *E. coli* exhibited stability regions across multiple levels of resolution (Figures 2C, 5C and 8C), including early “plateau” regions of considerable high stability that are not far from the “outbreak level” (where high instability was expectedly noticed). These “plateaux” represent specific genetic distance threshold ranges suitable for longitudinal monitoring of the main circulating lineages/genogroups, thus representing an important asset for the development/refinement of novel/existing nomenclature systems. This research line also informed us about the challenges associated with the identification of stability regions for *C. jejuni*, due to its high genetic diversity and polyclonal population structure ^78–80^. In fact, the identified regions for this species were often too short and less concordant between pipelines, anticipating great difficulties in the implementation of a robust hierarchical allele-based nomenclature system for this pathogen, and reinforcing the need for in-depth populational genomics studies to explore whether genogroup- or CC/ST-specific WGS-based classification systems could be an alternative solution to surpass these challenges. The identification of stability regions and their congruence with traditional typing have been the aim of multiple studies, typically relying on a single pipeline ^20,59,65^. In this study, we went a step further by performing this analysis for multiple pipelines. We observed not only that the threshold ranges with the highest congruence with traditional typing have a good overlap with stability regions, but also that different pipelines yielded similar results between them. For example, in *L. monocytogenes*, we could identify, in all pipelines, a high congruence with CC and ST classifications around cgMLST thresholds of 388-508 and 150-190 ADs, respectively. A similar scenario was found for *S. enterica*, where we observed moderate congruence between cg/wgMLST stability regions and serotype and ST classifications, roughly at 1261-1663 and 205-310 levels, respectively. The interpretation of these results should take into consideration the fact that our study relied on curated datasets that aimed at mimicking the current species diversity in public repositories, thus being likely biased towards more prevalent lineages and/or countries with high WGS volume. Still, the good inter-pipeline comparability and the high congruence with traditional typing are good indicators that our results are robust and can be extrapolated to other diverse datasets and scenarios. For example, the cg/wgMLST stability region that we identified as best representing serotype classification for *S. enterica* corresponded well with the largest stability region detected in a previous large-scale comparative genomics study of this species that relied on a different dataset ^65^. In summary, our investigation on cluster stability regions and congruence with traditional typing provides valuable results with possible implications for future nomenclature design, while favoring backwards compatibility with pre-WGS typing Era and increasing the confidence of laboratories to pursue the demanded technological transition.

Ultimately, with the rampant increase of WGS data and number of laboratories conducting WGS-based surveillance, we anticipate that the typical exercises for inter-laboratory comparability assessment (e.g. EQAs, ring trials, proficient tests, etc.), which are usually focused on outbreak cluster detection among a limited number of isolates, will need to evolve to another scale in terms of dataset size and diversity, and the magnitude of congruence assessment. In this regard, the methodologies and tools developed throughout this study may not only facilitate such inter-laboratory assessments, but also contribute to support the continuous intra-laboratory evaluation and long-term sustainability of existing or newly developed pipelines. In order to showcase this application, we employed the developed tools to rapidly assess the impact of modifying certain pipeline components (assembly and allele calling), showing that our results would be maintained if alternative assemblies have been used or if chewBBACA had been updated to a newer version.

In conclusion, this study provides valuable insights into the comparability of pipelines commonly used for FWD genomics surveillance and reinforces the need, while demonstrating the feasibility, of conducting continuous and comprehensive WGS pipeline comparison studies. Our work contributes to accelerate the technological transition towards a robust FWD WGS surveillance at global scale, while promoting a smoother communication and cooperation between One Health stakeholders.

## Methods

### Dataset selection and curation

In order to accomplish the objectives of this study, we aimed to compile a diverse dataset (including sequencing reads and respective metadata) that captures the genomic diversity within the populations of *L. monocytogenes*, *S. enterica*, *E. coli* and *C. jejuni*. To this end, the BeONE partners shared anonymized sequencing reads under a Material Transfer Agreement. Read QC, trimming and assembly were performed with Aquamis v1.3.9 ^95^ using default parameters. Briefly, reads were trimmed with fastp ^96^ and assembled with Shovill v1.1.0 ^97^ using SPAdes v3.15.4 *de novo* genome assembler ^98^. Assembly QC was performed with QUAST v5.0.2 ^99^, Augustus ^100^ for gene prediction and BUSCO ^101^. ConFindr v0.7.4 ^102^ was used for inter and intra genus contamination analysis and Kraken2 v2.1.2 ^103^ for read and assembly based taxonomy profiling. MLST ST determination was performed with mlst v2.22.0 ^21,104^. All genome assemblies passing the QC were included in the final dataset. Among the others, we noticed that a considerable proportion of assemblies was flagged as “QC fail” exclusively due to the “NumContamSNVs” parameter (252 out of 371 for *L. monocytogenes*, 198 out of 702 for *S. enterica*, 324 out of 675 for *E. coli* and 446 out of 919 for *C. jejuni*), suggesting that this setting might have been too strict. The manual inspection of a random subset of these samples showed that the detected contaminants were essentially related to sequencing errors. Therefore, we decided to recover the assemblies for which the percentage of reads corresponding to the correct species was >98% and integrate them in the final dataset. The final “BeONE dataset” comprised 1426 *L. monocytogenes*, 1540 *S. enterica*, 308 *E. coli* and 610 *C. jejuni* isolates ^47,49,51,53^. In order to ensure the dataset diversity, we complemented this “BeONE dataset” with publicly available WGS data for the four species of interest, and for which the QC and assembly followed the exact same methodology ^48,50,52,54,55^. The final dataset used in this study comprised a total of 3300 *L. monocytogenes*, 2974 *S. enterica*, 2307 *E. coli* and 3686 *C. jejuni* isolates.

### Pipeline and input selection

Based on the provided details for each pipeline, we established a strategy to avoid pipeline redundancy and increase the analytical scale (e.g., splitting the work between BeONE institutes with matching pipelines). As it would be unfeasible for each BeONE partner to filter the reads and/or assemble the genomes of so many isolates with the personal and computational resources available throughout the BeONE project timeframe, allele-based pipelines took as input all the assemblies available for these datasets, while SNP-based pipelines started from the QC-passed trimmed sequencing reads for the most represented STs/serotypes in each dataset, which were then aligned to the respective reference genome sequences (details in Table 1 and [Additional file](https://zenodo.org/records/12805751) 1). After an initial survey, we noticed that all allele-based approaches used by the BeONE partners that could be used to assemble the input datasets relied on SPAdes ^98,105^ for *de novo* genome assembly. Specifically, two non-commercial automated workflows were available within the consortium: AQUAMIS ^95,106^ and INNUca ^20,83^. As the use of different assembly pipelines was not expected to have a significant impact on the clustering results ^63^ and AQUAMIS turned out to be faster and consume less computational resources, we decided to use the assemblies generated with this tool (see “Dataset selection and curation”) as input for all allele-based pipelines. Of note, AQUAMIS is aligned with the QC and assembly workflow implemented in the EFSA One Health WGS analytical pipeline ^6,95^.

### Specific pipeline methodology

#### Allele-based pipelines

##### chewieSnake

The chewieSnake pipeline v3.1.1 ^17^ was used to perform allele calling with chewBBACA v2.0.12 (setting bsr_theshold: 0.6, size_threshold: 0.2) ^72^ on the genome assemblies, and to compute the distance matrix (grapetree_distance_method: 3) and generate a MST with GrapeTree v2.1 ^56^. Details on the used schemas are provided in [Additional file](https://zenodo.org/records/12805751) 38.

##### INNUENDO-like

The INNUENDO-like pipeline corresponds to a non-automated workflow similar to the one available at the INNUENDO platform ^20^. Allele-calling was performed with chewBBACA v2.8.5 (setting bsr_theshold: 0.6, size_threshold: 0.2) ^72^. Depending on the species, this pipeline was run with alternative schemas, as detailed in [Additional file](https://zenodo.org/records/12805751) 38. In order to distinguish the different approaches, except for *L. monocytogenes* for which only one schema was tested with this pipeline, for all the remaining species the pipeline name is followed by the schema used.

##### Ridom SeqSphere+

This software was run by two laboratories on different datasets. One was responsible for the analysis of *S. enterica* and *C. jejuni*, and the other one for the analysis of *L. monocytogenes* and *E. coli.* In both cases, SeqSphere+ version 8.3.4 (Ridom, Münster, Germany) was used to perform cgMLST allele calling on assemblies, and samples covering less than 95% of target loci of the schemes were removed from subsequent analysis. Details on the used schemas are provided in [Additional file](https://zenodo.org/records/12805751) 38. Of note, the partner Institute applying SeqSphere for *C. jejuni* typically run the cgMLST schema (637 loci) over closely related samples. A preliminary analysis revealed that the application of this methodology on the diverse *C. jejuni* dataset of this study would yield a very low clustering resolution, which hampered the comparison with the remaining ones. For this reason, an extra SeqSphere analysis with the extended cgMLST schema (637 core loci + 958 accessory loci) was requested to the partner in order to avoid the exclusion of this pipeline. The obtained wgMLST allelic matrix was subjected to clustering analysis with ReporTree v1.0.1 ^55^, as described below (this combined pipeline was designated SeqSphere-wgMLST). For the *S. enterica* dataset, hamming distances were calculated with SeqSphere+ version 8.3.4, and clustering analysis was performed with ReporTree v1.0.1 ^55^, as described below. For *L. monocytogenes* and *E. coli* datasets, hamming distances and HC single-linkage clustering were calculated in R with the hammingdists function of cultevo package v1.0.2 ^107^ and the hclust function of stats package v 0.1.0 ^108^, respectively.

##### Bionumerics

cgMLST analysis was performed on assemblies with BioNumerics 8.1 (Biomérieux). Details on the used schemas are provided in [Additional file](https://zenodo.org/records/12805751) 38. For all four target pathogens entries covering less than 95% of the loci in the scheme were removed. Entries with “multiple consensus loci” above 30 were removed. Furthermore, only sequences with genome sizes within the following size range were kept for further analysis: *L. monocytogenes* (2.8-3.1 Mb), *S. enterica* (4.5-5.3 Mb), *E. coli* (4.5-5.6 Mb) and *C. jejuni* (1.53-1.9 Mb). Hamming distances were also calculated with Bionumerics.

##### MentaLiST

cgMLST profiles were computed with MentaLiST v1.0.0 ^109^ (docker: mentalist:1.0.0--39e9e05e54) for each species of interest. MentaLiST ^109^ was used to build the *k*-mer databases for each species of interest with a kmer length (-k) of 31 and a minimum a percentage of allele coverage (-c) of 1.0, as well as call alleles from input fasta format (--fasta) with the original voting algorithm (--output_votes), and including alleles from ‘special cases’ such as incomplete coverage, novel, and multiple alleles (--output_special). Details on the used schemas are provided in [Additional file](https://zenodo.org/records/12805751) 38.

#### SNP-based pipelines

##### snippySnake (and WGSBAC)

snippySnake v1.1.0 ^41,110^ ran snippy v4.6.0 ^111^ on each sample followed by snippy-core to obtain the core-alignment and variant table. The snippy parameters were “mapqual: 60; basequal: 13; mincov: 10; minfrac: 0; minqual: 100; maxsoft: 10”. A SNP distance matrix was computed using snp-dists v0.7.0 ^112^. Details on the reference genomes are provided in [Additional file](https://zenodo.org/records/12805751) 1.

WGSBAC ^113^ ran snippy v4.6.0 ^111^ for each ST or serotype in standard settings for the sequencing data of each sample using the respective reference genome sequence (details in [Additional file](https://zenodo.org/records/12805751) 1). Based on core-genome SNP-alignments, snp-dists v0.6.3 ^112^ was used to calculate pairwise SNP-distances, which were used for hierarchical clustering with the hierClust function v.5.1 of the statistical language R ^108^. After the cluster congruence analysis for *L. monocytogenes*, it was observed that WGSBAC yields the exact same clustering results as SnippySnake. This was later confirmed for the other species (data not shown), so we only presented the results of SnippySnake, which also reflect the WGSBAC performance.

##### CSI Phylogeny

CSI Phylogeny v1.4 ^62^ was used to call SNPs and infer phylogenies. Settings used were: “min, SNP depth: 10”, “min relative SNP depth: 10%”, “Minimum distance between SNPs (prune): 10”, “min SNP quality: 30”, “min read map quality: 25”, “min Z-score: 1.96”, “ignore heterozygous SNPs: false”. In brief, the pipeline maps sequencing read data using BWA MEM^114^ to the chosen reference sequence (details in [Additional file](https://zenodo.org/records/12805751) 1), then vcfutils (part of SAMtools ^115^) is used to call SNPs. SNPs are then filtered by CSI Phylogeny ^62^, which produces a SNP matrix and a SNP pseudoalignment. Note that the usual CSI Phylogeny workflow would then produce a maximum likelihood tree from the alignment ^116^. However, in order to create comparable results between the methods, this final step was skipped in this study, and, instead, a single-linkage tree was created from the SNP matrix with ReporTree v1.0.1 ^55^, as described below.

##### SnapperDB

SnapperDB ^16^ software was utilized to assign a SNP address to each of the analyzed isolates. The ‘SNP address’ strain level nomenclature is used to describe the relationship between isolates in a defined population, and is routinely performed at the partner institute using hierarchical single linkage clustering at seven decreasing thresholds of genetic differences (250, 100, 50, 25, 10, 5 and 0 SNPs difference) to identify epidemiologically significant clusters. For the purpose of this study, partitions were identified at all possible SNP levels.

### Output harmonization and clustering at all resolution levels

For the cluster congruence analysis, it was important to harmonize the output of the different pipelines having clustering information at each possible distance threshold (i.e. all possible resolution levels). However, except for the WGSBAC pipeline and the Ridom SeqSphere+ pipeline for *L. monocytogenes and E. coli*, which provided a partitions table, the vast majority of them do not produce this type of output, but instead end up with outputs such as allele/SNP or distance matrices and phylogenetic trees (Table 1). Therefore, in order to have a harmonized input for the clustering congruence analysis, allele matrices (in the case of chewieSnake, INNUENDO-like, *C. jejuni* SeqSphere-wgMLST and MentaLiST) or distance matrices (in the case of Ridom SeqSphere+, Bionumerics, snippySnake, CSI Phylogeny and SnapperDB) were used as input for ReporTree v1.0.1 ^55^. This tool was used to process the available outputs and obtain clustering information at all possible distance thresholds. Default parameters were used, except for the argument “--loci-called”, which was set to 95% in order to remove samples with more than 5% missing loci, and for the argument “--site-inclusion”, which was set to 99% for the INNUENDO-like using the INNUENDO wgMLST schemas and the *C. jejuni* SeqSphere-wgMLST pipeline in order to only keep informative wgMLST loci with alleles assigned for 95% of the samples ([Additional file](https://zenodo.org/records/12805751) 38). ReporTree was run for all of them using the single-linkage hierarchical clustering algorithm, which is also the default clustering method that the respective institutes use to determine clusters of potential public health interest. For all cases where allelic matrices were available, an additional ReporTree run was performed requesting clustering with the MSTreeV2 GrapeTree algorithm ^6,24,56,64^.

### Traditional typing

The traditional typing information used in this work corresponded to the ST and CC for *L. monocytogenes*, ST and serotype for *S. enterica* and *E. coli*, and ST and CC for *C. jejuni*. ST was determined for all species with mlst v2.22.0 ^21,104^ through Aquamis v1.3.9 ^95^, as described above (“Dataset selection and curation” section). *L. monocytogenes* and *C. jejuni* CCs were inferred for each predicted ST based on the information present in PubMLST/BIGSdb ^21^ schema profiles for this species. *S. enterica* serotype was determined with SeqSero2 v1.1.1^117^ using default parameters. *E. coli* serotype was determined for QC-passed reads with patho_typing v1.0 ^118^ using default parameters.

### PopPUNK

The main genomic lineages present in *E. coli* and *C. jejuni* datasets were inferred with PopPUNK v2.6.5 (default parameters) using *E. coli* v2 and *C. jejuni* v1 databases obtained on November 22^nd^, 2023 from BacPop website (https://www.bacpop.org/poppunk/ ^119^) and providing AQUAMIS genome assemblies as input. Given the unavailability of functional databases for *L. monocytogenes* and *S. enterica* at the time of this study, this tool was not used for these two species.

### Identification of pipeline stability regions

Stability regions, i.e. distance thresholds providing similar clustering results were determined with ReporTree v1.0.1 ^55^ using default parameters. Briefly, this pipeline uses the script *comparing_partitions_v2.py* ^55,120^ to assess the number and composition of clusters obtained at progressively increasing distance thresholds and determine the neighborhood Adjusted Wallace coefficient (nAWC) at consecutive partitions (“n + 1” → “n”), based on a previously described approach ^57–59^. This script was run with default settings requesting the “stability” analysis, and, therefore, a region was considered as of stability when a nAWC ≥ 0.99 was observed in more than 5 subsequent thresholds.

### Congruence analysis

Cluster congruence was assessed with the script *comparing_partitions_v2.py* ^55,120^ requesting the “between_methods” analysis. Briefly, for each pairwise comparison, we calculated the AWC, as a measure of the probability that two samples that cluster together using one method (at a given threshold level) also cluster together with another one (at a given threshold level) or belong to the same lineage, ST, CC or serotype ^58^. This was conducted between all possible threshold levels in both directions (method A → method B and method B → method A). In addition, for each comparison, we also calculated the Adjusted Rand (AR) coefficient as a measure of the overall agreement between the typing methods ^58^. The three values calculated for each comparison were then combined into a “Congruence Score” (CS) (CS = AWC_A → B_ + AWC_B → A_ + AR), which varies from 0 (no congruence) to 3 (absolute congruence). This score was used to compare clustering results at each possible threshold of a pipeline either with traditional typing data or with each of the possible thresholds of another pipeline.

### Identification of correspondence points

The script *get_best_part_correspondence.py* ^121^ was used for each pipeline comparison in order to assess what was the threshold that provided the most similar clustering results in the other pipeline (i.e., the best “correspondence point”), as assessed by CS scores. Only comparisons yielding CS ≥ 2.85, which ensures a score ≥ 0.95 for each CS metric component, were considered as possible “correspondence points” (i.e., no correspondence was reported when the best match had CS < 2.85). When a given threshold had a single valid correspondence point, this was assumed as the point of highest congruence. When a given threshold had several valid correspondence points, the closest threshold was reported as the best match. The trendline fitting the correspondence points of each comparison, as well as the respective r^2^ and slope, were determined with the scipy v1.10.0 linregress function ^122^.

### Assessment of the discriminatory power at high-resolution thresholds

The script *comparison_outbreak_level.py* ^121^ was used to assess the discriminatory power of each pipeline at high-resolution thresholds. Briefly, all the clusters identified at a potential outbreak level (7 ADs for *L. monocytogenes*, 14 ADs for *S. enterica*, 9 ADs for *E. coli*, and 4 ADs for *C. jejuni*, see section “Thresholds for identification of outbreak clusters” for details) by each allele-based pipeline and their composition was recorded. Then, a list with the union of these clusters was created and the lowest threshold level at which each cluster was identified with the same composition in each pipeline (allele- and SNP-based) was assessed. Only samples passing the QC of all pipelines were considered for these analyses.

The script *stats_outbreak_analysis.py* was used to determine the number of clusters identified by a given pipeline at a given threshold that could also be detected with the exact same composition by another pipeline at a (or up to a) similar or even higher threshold. Only samples passing the QC of all pipelines were considered for these analyses. Clusters with overlapping samples but different compositions were considered as different clusters.

Regarding the assessment of the genetic diversity within potential outbreak-related clusters, we used the script *stats_outbreak_analysis_snp_dists.py*. This script determines, for each cluster identified by a given allele-based pipeline at a given threshold, the maximum allelic or SNP distance within the cluster that the same or an alternative allele- or SNP-based pipeline detected with the exact same composition.

### Thresholds for identification of outbreak clusters

The threshold used for identification of potential outbreak clusters varied between species. For *L. monocytogenes*, we used a cutoff corresponding to 7 ADs, as this is the threshold conventionally used to determine potential outbreak-related samples in this species ^7,63,123^. For the other three, namely *S. enterica*, *E. coli* and *C. jejuni*, we considered a dynamic approach using the level for outbreak detection determined by in the INNUENDO project ^20^. Specifically, 0.43% of the core loci for *S. enterica*, 0.34% for *E. coli*, and 0.59% for *C. jejuni*. For *S. enterica* and *E. coli,* the corresponding absolute threshold translated to 14 and 9 ADs in all pipelines, respectively, which are absolute thresholds similar to the ones obtained in INNUENDO ^20^. For *C. jejuni*, the calculated absolute threshold varied between pipelines, corresponding to 4 ADs for the pipelines running shorter schemas and 8 ADs for those running the PubMLST schema. However, as previous studies suggested the application of a 4 ADs threshold also for the PubMLST schema ^81,82^, we decided to run our exercise with a 4 ADs threshold in all pipelines. Noteworthy, in order to provide more informative results regarding the inter-pipeline pipeline comparison at outbreak level, this study also explored other commonly used thresholds (e.g. 4 ADs in *L. monocytogenes* and 5 ADs in *S. enterica*). Additionally, taking advantage of the flexibility of *stats_outbreak_analysis.py* (described above), the exercise included the assessment of the proportion of clusters identified by a given pipeline at a static threshold (= AD1) that could also be detected with the exact same composition when: i) a static threshold (= AD2) is also applied to the second pipeline (e.g. Figures XC, YC, WC and ZC); or ii) a range of thresholds (≤ AD2) is also applied to the second pipeline (e.g. Figures XD, YD and WD).

### wgMLST zoom-in exercise

For the three species with wgMLST schemas available, namely *S. enterica*, *E. coli* and *C. jejuni*, we conducted an additional exercise that aimed to assess the potential of a dynamic cgMLST approach to increase the discriminatory power at outbreak level. Briefly, for each of the three species, the INNUENDO-like pipeline was run with the respective INNUENDO wgMLST schema ([Additional file](https://zenodo.org/records/12805751) 38) and ReporTree v1.0.1 ^55^ was used to identify clusters at all possible distance thresholds, as described above. For the purpose of this exercise, the “--zoom-cluster-of-interest” argument of ReporTree was set to the threshold used for outbreak identification (see section “Thresholds for identification of outbreak clusters”). With this approach, for each cluster identified at this threshold level, the set of core loci was dynamically increased according to the samples that belong to the cluster. The maximum AD difference between two samples within a cluster and the difference between this value and the maximum distance observed in the initial analysis (i.e. with the cgMLST loci determined for the whole dataset) for the same cluster was determined with the script *wgmlst_exercise.py*. Of note, in *C. jejuni*, a similar analysis was performed for the SeqSphere-wgMLST pipeline. Moreover, as the INNUENDO-like pipeline using the INNUENDO static cgMLST schema was the one providing the lowest resolution in *C. jejuni*, for this species, we also tested the impact of performing a dynamic cgMLST approach using the INNUENDO wgMLST schema in outbreak clusters identified in an initial analysis with the static INNUENDO cgMLST schema. For this, we performed an additional ReporTree run, where, instead of providing the initial allele matrix for the static cgMLST, we provided the allele matrix for the wgMLST schema. Moreover, the file combining metadata and the clustering partitions at all levels that was obtained in the initial ReporTree run (with the static cgMLST schema) was provided as metadata input. This additional ReporTree run was performed for each cluster identified in the initial run at the threshold used for identification of outbreak clusters (i.e. 4 ADs) combining the “--filter” and “--subset” arguments.

### Assessing the impact of modifying pipeline components

This study involved the distribution of the same set of genome assemblies (generated with AQUAMIS ^95^) across multiple institutes in order to assess inter-pipeline cluster congruence. This approach was taken because it would be unfeasible for every BeONE partner to run their own assemblies with the personal and computational resources available throughout the BeONE project timeframe, and also because the use of different assembly pipelines was not expected to have a significant impact on the clustering results ^63^. Nevertheless, in order to assess the impact of providing genome assemblies performed with AQUAMIS ^95^ in a pipeline that does not use this assembly workflow, and of a possible future update of chewBBACA ^72^ allele-caller, we performed two additional analysis: i) comparison of the clustering results obtained with same pipeline when AQUAMIS or INNUca ^20,83^ assemblies are provided as input; and ii) comparison of the clustering results obtained with the same pipeline when two different versions of chewBBACA are used. For the first analysis, we assembled the datasets of each of the four species with INNUca v4.2.2 ^20,83^ with default parameters. Afterwards, the INNUENDO-like pipeline (with chewBBACA v2.8.5) was run in parallel using as input the INNUca and the AQUAMIS genome assemblies of the set of samples that passed the QC of the two assembly workflows. For the second analysis, for each of the four species, we ran in parallel the INNUENDO-like pipeline over the AQUAMIS genome assemblies using chewBBACA v2.8.5 and chewBBACA v3.3.5 with the non-populated cgMLST schemas available on April 1^st^, 2024 in chewie-NS ^75,124^. After an initial run of the allele-caller to identify the samples with less than 5% of missing loci, we then run the two versions of the allele-caller on the set of passing samples. For each of the two comparisons (different assembly workflows and different allele-caller versions), we performed a cluster congruence analysis and an in-depth pairwise comparison at outbreak level following the above-mentioned methodologies.

### Graphical visualization

Plots generated in this research study were generated with the Seaborn python module ^125^ or with the ggplot2 R package ^126^.

### Data availability

Anonymized sequencing reads of the “BeONE dataset” are deposited in the European Nucleotide Archive (ENA) database under the BioProjects PRJEB57166, PRJEB57179, PRJEB57098 and PRJEB57119. Genome assemblies are deposited in the Zenodo repository (*L. monocytogenes*: 10.5281/ZENODO.7267486 ^47^; *S. enterica*: 10.5281/ZENODO.7267785 ^49^; *E. coli*: 10.5281/ZENODO.7267844 ^51^; *C. jejuni*: 10.5281/ZENODO.7267879 ^53^). The public dataset data was retrieved from Zenodo (*L. monocytogenes*: 10.5281/ZENODO.7116878 ^48^; *S. enterica*: 10.5281/ZENODO.7119735 ^50^; *E. coli*: 10.5281/ZENODO.7120057 ^52^; *C. jejuni*: 10.5281/ZENODO.7120166 ^54^). The collection of scripts used to conduct these analyses are available at the github repository https://github.com/insapathogenomics/WGS_cluster_congruence ^121^. Supplementary data are available in the Zenodo repository (https://doi.org/10.5281/zenodo.12805750) ^133^.

## Funding

This work was supported by co-funding from the European Union’s Horizon 2020 Research and Innovation program under grant agreement No 773830: One Health European Joint Programme (2020–2022) (https://onehealthejp.eu/projects/foodborne-zoonoses/jrp-beone) and by the ISIDORe project (funding from the European Union’s Horizon Europe Research & Innovation Programme, Grant Agreement no. 101046133). VM contribution was funded by national funds through FCT - Foundation for Science and Technology, I.P., in the frame of Individual CEEC 2022.00851.CEECIND/CP1748/CT0001 (2023 onwards). JDS contribution was supported by the project “Sustainable use and integration of enhanced infrastructure into routine genome-based surveillance and outbreak investigation activities in Portugal’’ (GENEO, https://www.insa.min-saude.pt/category/projectos/geneo/) on behalf of the EU4H programme (EU4H-2022-DGA-MS-IBA-1). Research at the National Veterinary Research Institute (PIWet) Poland was supported by the Polish Ministry of Education and Science from the funds for science in the years 2018-2022 allocated for the implementation of a co-financed international project.

## Data Availability

Anonymized sequencing reads of the BeONE dataset are deposited in the European Nucleotide Archive (ENA) database under the BioProjects PRJEB57166, PRJEB57179, PRJEB57098 and PRJEB57119. Genome assemblies are deposited in the Zenodo repository (L. monocytogenes: 10.5281/ZENODO.7267486; S. enterica: 10.5281/ZENODO.7267785; E. coli: 10.5281/ZENODO.7267844; C. jejuni: 10.5281/ZENODO.7267879). The public dataset data was retrieved from Zenodo (L. monocytogenes: 10.5281/ZENODO.7116878; S. enterica: 10.5281/ZENODO.7119735; E. coli: 10.5281/ZENODO.7120057; C. jejuni: 10.5281/ZENODO.7120166). The collection of scripts used to conduct these analyses are available at the github repository https://github.com/insapathogenomics/WGS_cluster_congruence. Supplementary data are available in the Zenodo repository (https://doi.org/10.5281/zenodo.12805750).

https://doi.org/10.5281/ZENODO.7267486

https://doi.org/10.5281/ZENODO.7116878

https://doi.org/10.5281/ZENODO.7267785

https://doi.org/10.5281/ZENODO.7119735

https://doi.org/10.5281/ZENODO.7267844

https://doi.org/10.5281/ZENODO.7120057

https://doi.org/10.5281/ZENODO.7267879

https://doi.org/10.5281/ZENODO.7120166

## Acknowledgements

The authors thank INSA’s National Reference Laboratory for Gastrointestinal Infections, headed by Dr. Mónica Oleastro, and Dr. Magdalena Zając and Prof. Dariusz Wasyl from PIWET’s Department of Microbiology, National Veterinary Research Institute for sharing WGS sequencing data of foodborne bacterial isolates. The authors also thank Dr. João André Carriço for the productive discussions throughout this research study. The authors also recognize the contribution of the National Distributed Computing Infrastructure of Portugal (INCD) and the Norwegian Research Infrastructure Services (NRIS) for providing the necessary resources to run the genome assemblies. INCD was funded by FCT and FEDER under the project 22153-01/SAICT/2016. We also thank the Italian Ministry of Health for supporting the acquisition of high-performance computing resources. This publication made use of the PubMLST website (https://pubmlst.org/) developed by Keith Jolley and sited at the University of Oxford. The development of that website was funded by the Wellcome Trust.

## Conflict of interests

The authors declare no conflict of interests.

## Notes

### Competing Interest Statement

The authors have declared no competing interest.

